# *Nowcast-It:* A Practical Toolbox for Real-Time Adjustment of Reporting Delays in Epidemic Surveillance

**DOI:** 10.1101/2025.08.19.25333998

**Authors:** Amna Tariq, Ping Yan, Amanda Bleichrodt, Gerardo Chowell

## Abstract

Reporting delays caused by delays in case detection, symptom onset after infection, seeking medical care, or diagnostics distort the accurate forecasting of diseases during epidemics and pandemics. This inherent delay between the time of symptom onset and the time a case is reported is known as the reporting delay. To address this, we introduce a practical nowcasting approach grounded in survival analysis and actuarial science, explicitly allowing for non-stationarity in reporting delay patterns to better capture real-world variability. Despite its relevance, no flexible and accessible toolbox currently exists for non-stationary delay adjustment. Here, we present Nowcast-It, a tutorial-based toolbox that includes three components: (1) an R codebase for delay adjustment, (2) MATLAB algorithms for performance evaluation, and (3) a user-friendly R-Shiny application to enable interactive visualization and reporting delay correction without requiring coding expertise. The toolbox supports daily, weekly, or monthly resolution data and enables model performance assessment using metrics such as mean absolute error, mean squared error, and 95% prediction interval coverage. We demonstrate the utility of *Nowcast-It* using publicly available weekly Ebola case data from the Democratic Republic of Congo. We and others have adjusted for reporting delays in real-time analyses (e.g., Singapore) and produced early COVID-19 forecasts; here we package those delay-adjustment routines into an accessible toolbox. It is designed for researchers, students, and policymakers alike, offering a scalable and accessible solution for addressing reporting delays during outbreaks.

## Introduction

The timely and accurate maintenance of disease surveillance systems helps to provide prompt information required to identify risks, predict challenges, allocate public health resources, monitor disease trends, and deploy interventions to contain outbreak situations such as the COVID-19 pandemic [1]. However, despite the importance of timely surveillance, delays in reporting case information exist in real time [2] often because the reporting of symptomatic cases is subject to reporting delay, which is defined as the inherent delay between the time of symptom onset and the time a case is reported in the surveillance system. These delays can vary widely depending on the disease and the health system, ranging from several days (e.g., for influenza and Ebola) to months (e.g., for tuberculosis), and even years in the case of AIDS. Indeed, reporting delays stem from a mix of biological, behavioral, and operational factors. These include various controllable and uncontrollable processes, such as the incubation period of the infection, delay in case detection, delay in symptom of onset after infection, delay in seeking medical care, delay in diagnostics, delay in processing of surveillance systems, non-contacts, community deaths, and the time to report the case in the database etc. [8, 9]. Furthermore, a strong day-of-the-week effect (i.e., very few cases are reported on Saturdays and Sundays) often influences the reporting of cases to surveillance systems [3]. Reporting delays are also enhanced in resource-constrained settings, resulting in the number of new cases by the time of symptom onset showing a downward bias towards the current day [3] [4].

Nowcasting is the process of estimating the actual number of symptomatic-but-not-yet-reported cases in real time, which is essential for correcting this bias and enhancing situational awareness during an outbreak. A range of nowcasting models has been proposed to address this challenge. Bayesian smoothing approaches such as NobBS [2] leverage historical patterns and temporal correlation between incidence values to generate probabilistic nowcasts, while constrained P-spline smoothing methods [3] model the reporting intensity surface across onset and delay dimensions. These methods offer valuable flexibility and uncertainty quantification but often assume stationarity in reporting delays and can be computationally intensive or require substantial statistical expertise. In this tutorial paper, we introduce Nowcast-It, a user-friendly toolbox based on a frequentist approach rooted in survival analysis and actuarial science. The toolbox estimates reporting delay-adjusted incidence using reverse-time hazard methods and allows for non-stationary delay distributions by specifying a moving window for delay probability estimation. The underlying delay-adjustment routines have been used in previous real-time analyses during outbreaks, including COVID-19 [1, 5] and Ebola [4]. The reporting delay-adjusted case incidence estimates can then be utilized to fit and forecast trajectories of infectious diseases using phenomenological dynamic growth models or mechanistic models. This toolbox is written for various audiences, including students training in infectious disease modeling, emergency preparedness and pandemic control, epidemiological time-series forecasting, and dynamic growth modeling. The toolbox is also useful for researchers and policymakers who need to conduct short-term forecasts by relying on historical and real-time trajectory data of the process of interest, such as an unfolding epidemic. We provide MATLAB code to assess the performance metrics to choose the best window for non-stationary reporting delays. We have also developed an R-Shiny application to simplify the usage of the toolbox further.

Unlike popular statistical nowcasting techniques that principally draw from survival analysis and actuarial methods to model the reporting delays, assuming stationarity (quasi-stationarity) of the reporting delays, our approach uses a flexible, windowed estimation of the delay distribution to accommodate shifts due to changing surveillance systems or field conditions. This makes the toolbox particularly well-suited for low-resource settings or emergencies where surveillance patterns evolve over time [6].This robust method employs survival analysis techniques and uses point estimation based on reverse time hazards [7]. The 95% prediction limits are derived using the statistical estimations introduced by Lawless, modified by assuming non-stationary reporting delay probabilities [8, 9]. The toolbox includes the creation of line list data, estimation of reporting delay-adjusted trends, variable reporting delay window, and ad-hoc approaches to adjust for reporting delays.

This tutorial-based primer is organized as follows. After providing an overview of the toolbox functions for users, we first introduce the reporting delay adjustment method included in the toolbox, and then describe the underlying methodology, user parameters, and functions to adjust for the reporting delay. Finally, we use specific examples in the context of the Ebola epidemic in the DRC to demonstrate the functions for generating, displaying, and quantifying the performance of nowcasts. A description of the shiny app’s usage follows towards the end. A tutorial video demonstrating the toolbox functionality is available at the you tube link: https://www.youtube.com/watch?v=KDYAnXb-IEY&ab_channel=Chowell_Lab.

## Methods

### Implementation

#### Installing the toolbox

Download the R code located in the ‘**Reporting delay adjustment code**’ folder from the GitHub repository: https://github.com/atariq2891

Create an ‘input’ folder in your working directory where your input data will be stored.

- Create an ‘output’ folder in your working directory where the output files will be stored.
- Create a ‘Functions’ folder in your working directory where the function files will be stored.
- Download and install R studio.
- Open an R session so a window called R console appears.
- Install and run xQuartz (XQuartz-2.8.5.pkg) from https://www.xquartz.org/ to visualize the figures produced by the reporting delay adjustment algorithm.

For the performance metrics

- Download MATLAB

#### Overview of the toolbox functions

Table 1 lists the names of user functions associated with the toolbox along with a brief description of their role.

**Table 1:** Description of user functions associated with the toolbox.

| Function | Type | Role |
| --- | --- | --- |
| datainput.txt | User | Reads a data file as input matrix D |
| ddelay.txt | User | Contains the algorithms of lawless based on reverse time hazards and survival analysis. The input data is the matrix "mat". There is a second choice, called "m". This is to select the most recent "m" weeks (according to week of reporting) that we believe the reporting practice has been reasonably stable |
| delay1.txt |  | Creates a temporary working matrix to be used in ddelay.txt function |
| matrixfill.txt | User | Creates the matrices for the right-truncated data |
| psum.txt | User | Adds the components of the matrices |
| shift.txt | User | Shifts the vector to the right or left |
| shiftvec.txt | User | Creates the time lagged matrix. Shifts the vector to the right or left |
| strip.txt | User | If else function for the matrices |

#### Data resolution

The case or mortality data utilized in the nowcasting methodology can be based on a daily, weekly or monthly resolution, depending on the availability and noise in the data.

### Reporting delay adjustment methodology

#### Nowcasting approach

For this tutorial, we have developed a method to adjust the epidemic curve for the reporting delays utilizing the Actuaries’ method suggested initially by Taylor. G in 1986, where it is known as the “claims reserving modeling” method [7]. This method found its application in the biostatistical context analyzing the HIV AIDS data [8, 10]. Since then, this method has been widely utilized to understand the reporting delays in the context of various diseases such as the Ebola virus disease and COVID-19 [1, 11]. We address the nowcasting approach based on the Actuaries’ method in the statistical framework of the occurred but not reported events [8]. Here, the estimation of the delay distribution takes into account the inherent right-truncation of the data-generating process. In general, this method employs survival analysis techniques and uses point estimation based on reverse time hazards [7]. The 95% prediction limits are derived using the statistical estimations introduced by Lawless, modified by assuming non-stationary reporting delay probabilities [8, 9]. Daily, weekly or monthly data can be utilized to ascertain the reporting delays. However, mostly weekly time intervals are used as a compromise between maximizing the temporal resolution and reporting irregularities in batch reporting of daily counts and inaccuracies in retrospectively ascertained dates of onset.

To explain the approach, we will utilize the weekly Ebola data from the 2018 Ebola epidemic in the DRC. The discrete time series data published by the week of reporting provides the Ebola case counts by the week of symptom onset. In this time series data, each reported individual is associated with two events; the first event is directly related to the time of illness onset, and the second event is directly related to the diagnosis of the case. Let *C* denote the “current time,” which is the time point on which data are used for analysis. Let *t* = 0,1,2 … *C* denote the time of the occurrence of the event; *t* = 0 is the earliest possible time when an event can occur in the population. Let *x* = 0,1,2, …. *C - t* denote the reporting delays, where *x* = 0 indicates that the event is reported in the same time-period when it occurs. Therefore, the data can be grouped as

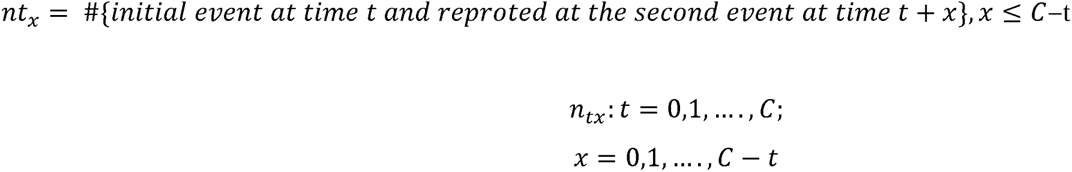

The row totals can be denoted as : 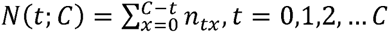

The column totals can be denoted by: 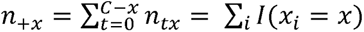

This data only fills the upper triangular array of the matrix due to right truncation, with column totals representing the number of events with *X = x* and row totals representing the number of events as reported by the current time, *C* (Fig 1).

**Fig 1.**
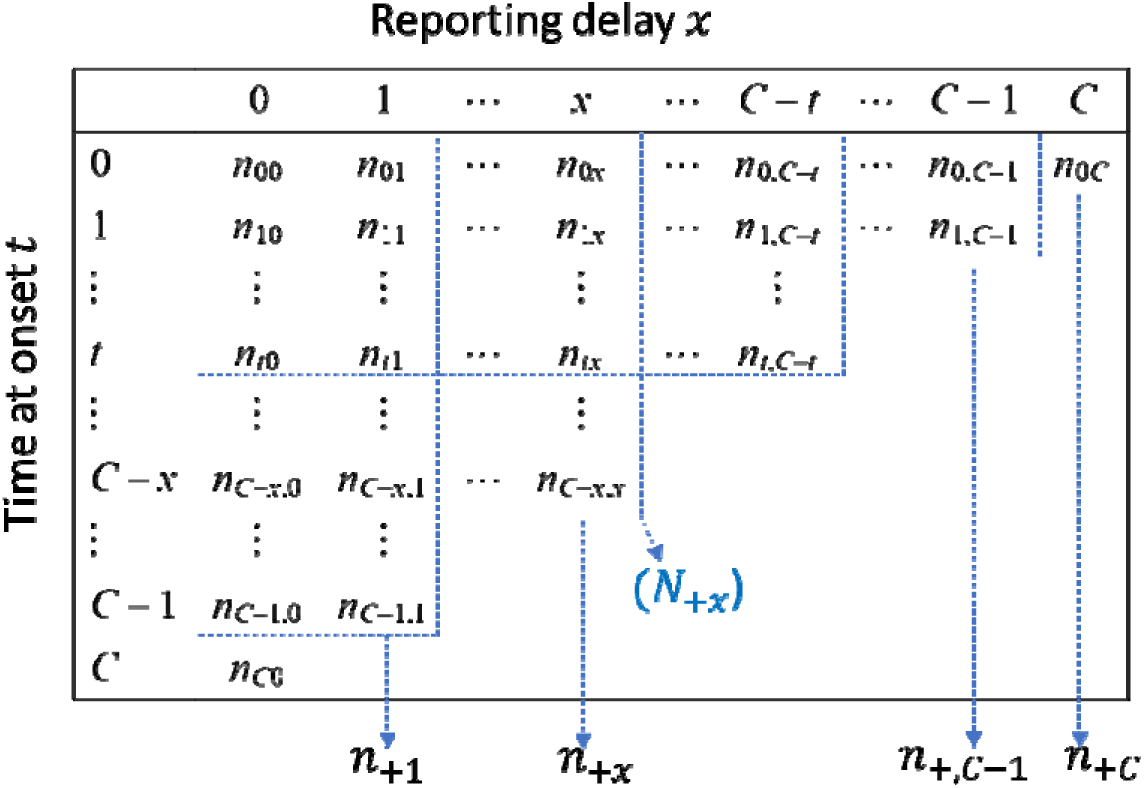
Upper triangular matrix depicting the reporting delays.

We define an estimate as  ___    where       .     It means that    _______ out of events in    .   This follows;         =    _______  ______  _______,   where       This means that the proportion of events with delay      out of those with delay  .

The reporting delay adjustments are based on the above-mentioned reverse hazard function and given as   _____.

Hence, the reporting delay adjustment factor assuming stationarity (also known as quasi-stationarity) of the reporting delays patterns can be described as;     ___________

Where,  estimates the proportion of individuals with time onset at time reported by time,  out of those would be reported by time  and  is the reporting delay adjustment. Assuming stationarity across the reporting delay patterns yields less robust estimates of delay adjustment, we use a modified approach. We choose the most recent reporting periods (assuming relatively stable reporting delay patterns in the most recent reporting time periods), owing to the irregular reporting delay patterns captured by the surveillance systems over longer periods.

#### Assuming non-stationarity in the reporting delays

Since reporting delays are not stationary in the real world, we employ a modified approach by selecting a variable reporting delay window, defined as “m”, to address this issue of stationarity. Disease reporting patterns are generally influenced by external factors (i.e., delay in disease detection, late diagnostic test) and are unstable. Since we observe a systematic departure from stationarity of the reporting delays in the data used for the tutorial (because of change of reporting system and observable events), we treat reporting patterns to be stable during the different smaller reporting periods (like the last 6 weeks or last 8 weeks of the data), implying non-stationarity during those periods only. Hence, this window “m” represents the trends of the reporting delay probabilities for the chosen time-period, assuming stable reporting delay patterns in that time-period. We use different values of “m” and compare the performance of different “m’s” across the data sets, each set apart by one week. The “m’s” employed in this study include the most recent reporting periods (7,9 and 11 weeks, and 3,6 and 8 weeks) along with the best “m” value. For choosing the best “m” value we plot the ratio of delays that occur in the same week (delay 0) to the delays that occur in the first week of onset (delay1) (ratio delay0/delay1). This provides us with the calculated probability for the last prediction. This one-step prediction can be quantified as follows:

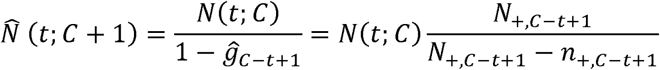

However, it can only be performed for *t* = 1, 2, …, *C* because *C*_l_ ≤ *C* = *t*. In this equation, 1 - ĝ_*C-t*+l_ estimates the proportion of individuals with onset at time *t* reported by time *C*, out of those who would be reported by time *C*_l_ = *C* + 1. The performance of this one-step prediction (also known as the partial delay adjustment) also informs us about the performance of our ultimate reporting delay adjustment. Sometimes we use the reporting delay distribution pattern for the entire data set (assuming a stationary reporting delay distribution across the entiree dataset) to compare the differences between choosing stable versus variable reporting delay patterns that are best suited for a data set to achieve the most robust adjustment of the raw incidence.

#### Evaluating the performance of nowcasts, performance metrics

To assess the performance of our nowcasting method and the nowcasts produced using differing values of “m”, we employ three performance metrics: mean absolute error (MAE), mean square error (MSE), and the 95% prediction interval. The MAE is given by

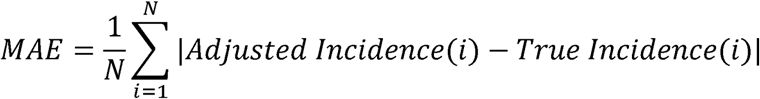

and the MSE is given by

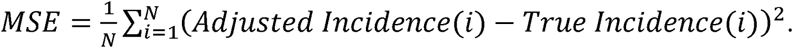

In addition, we assessed the coverage of the 95% prediction interval, the proportion of the observations that fell within the 95% prediction interval. The 95% prediction coverage is given by:

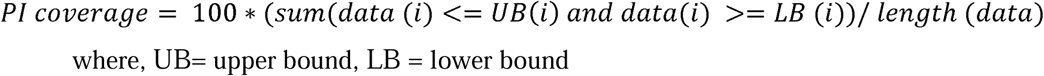

In this tutorial example, the performance metrics are estimated against the true incidence reported 8 weeks later (*n* - 8) for the same dates of onset. We use Maltab R-2023a Inc to estimate the performance metrics.

## Results and Discussion

### The input dataset

For this toolbox, the time series data must be stored as line-listed data in the ‘input’ folder and needs to be a text file with the extension *\*.txt*. Data can be stored in the form of case or death count data. For this tutorial purpose we are using case count data. Each case should be a separate row in the data set, such that the first column should correspond to the date/week of symptom onset: 1,2,3, …, and the second column corresponds to the date/week of reporting: 1,2,3 If you are using the dates of onset and reporting, the five-digit number representing the date of onset, counted from 04/30/2018, and the date of reporting, 08/20/2018, is created by selecting two fields originally entered in date format and converting them to number format in Excel. For example, 08/20/2018 is converted to 43332.00. This Excel file should then be saved as a *.txt* file.

To illustrate the methodology presented in this tutorial paper, we employ the weekly incidence curve of the Ebola epidemic in the DRC. Data is extracted from the situation reports publicly available from the World Health Organization website [12]. The data file is located in the input folder within the working directory (data file path: ./input/March_3_2019.txt). A snapshot in Excel (Fig 2) and *.txt* (Fig 3) of the contents of the file is shown below:

**Fig 2.**
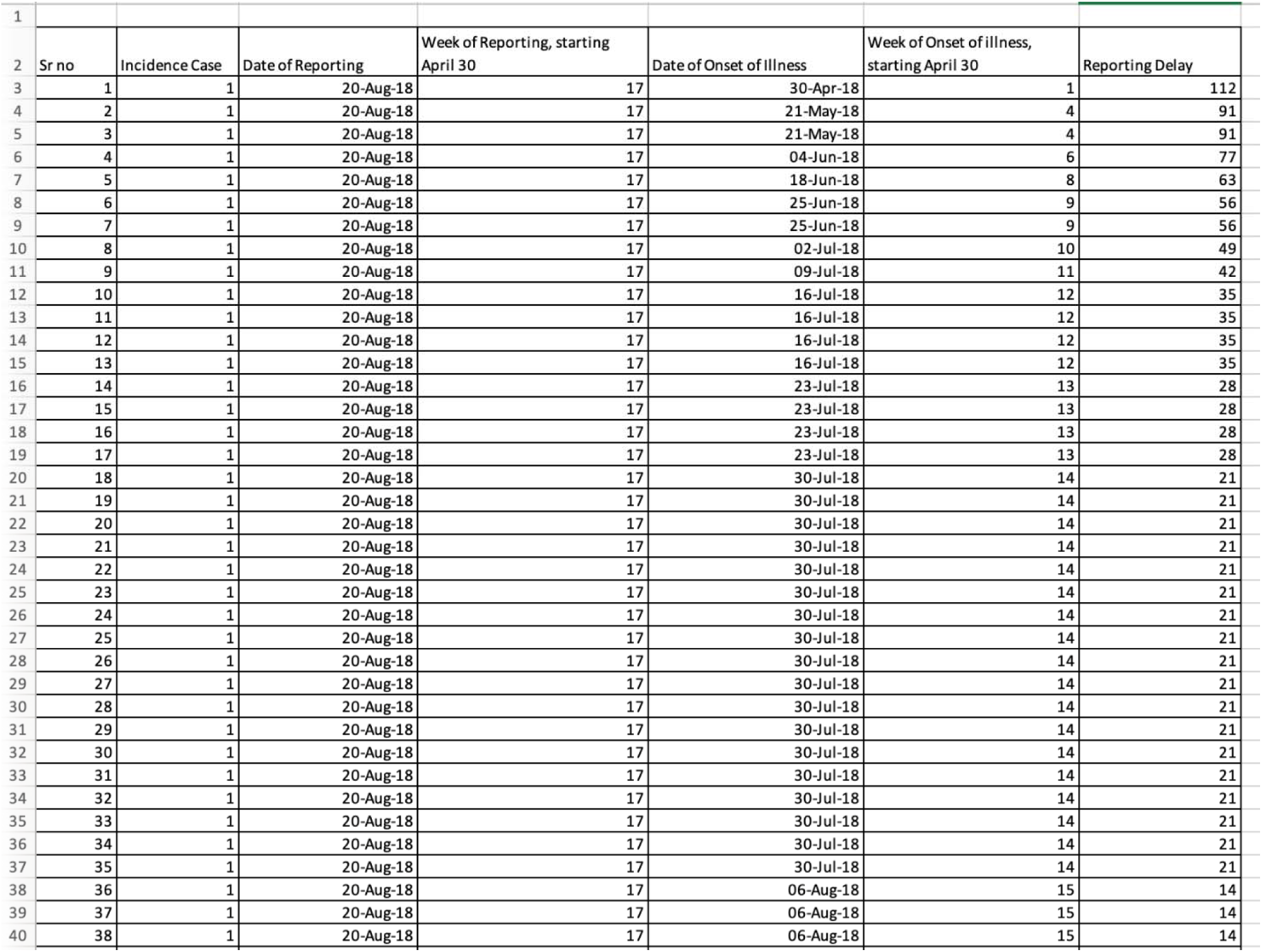
Snap-shot of the excel data file created from the WHO situation report published on March 3, 2019.

**Fig 3.**
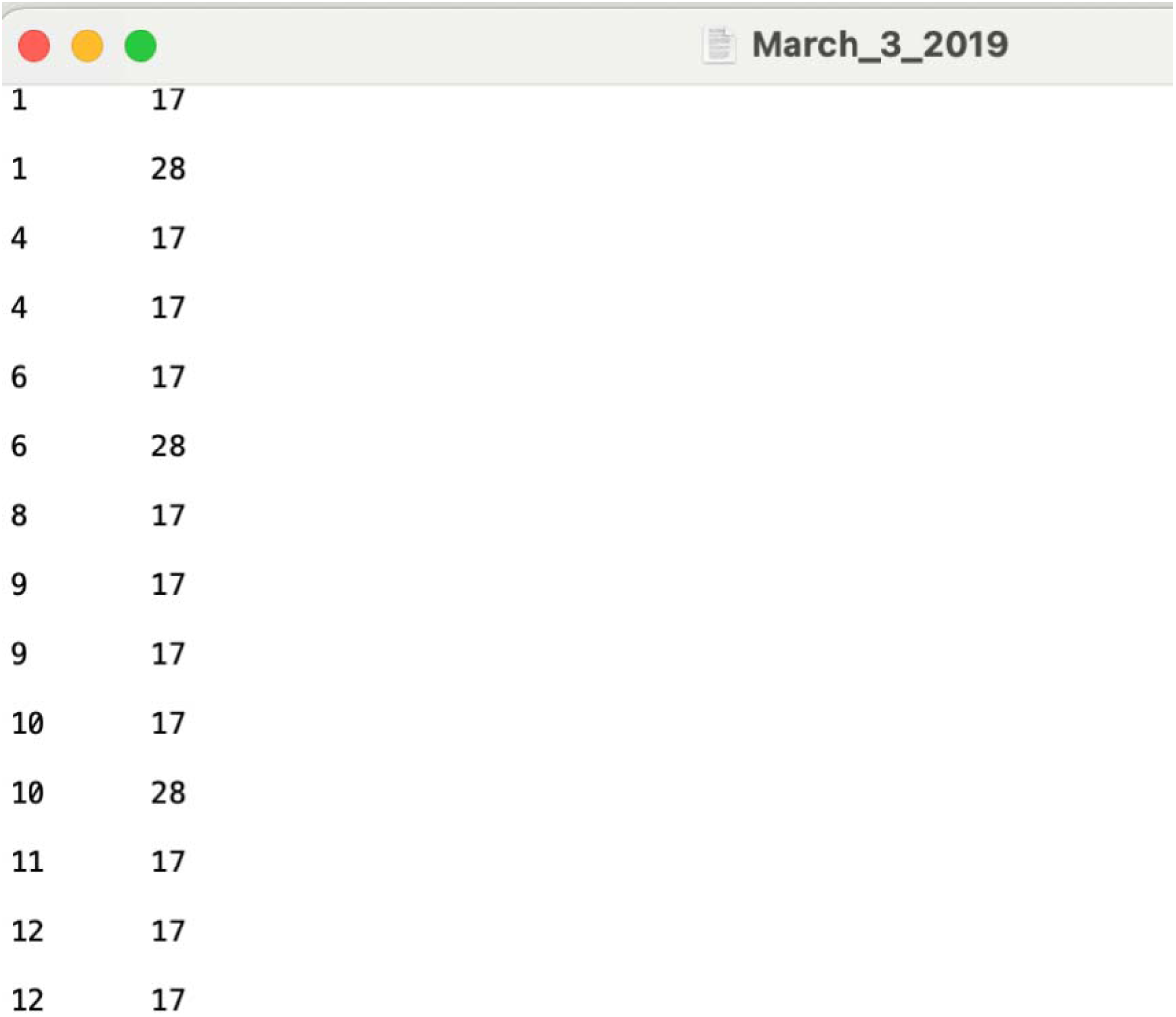
Snap-shot of the .txt file saved in the number format for the excel file dated on March 3, 2019. Dat is stored at a weekly resolution. The first column represents the week of symptom onset, and the second column represents the week of reporting of the Ebola cases.

The *load-data-and-functions.R* file is an RStudio program that allows users to load all functions into the algorithm (Fig 4). This file also contains the algorithm to produce a 2-way crosstabulation of case/death counts that are generated as mat0, as structural zero’s fill in the matrix. This matrix should be a square upper-right triangle matrix representing the week of onset versus the week of reporting. The algorithm also produces an upper-left triangular square matrix, mat, representing the week of onset versus the week of reporting. This matrix is utilized in the “ddelay” function.

**Fig 4.**
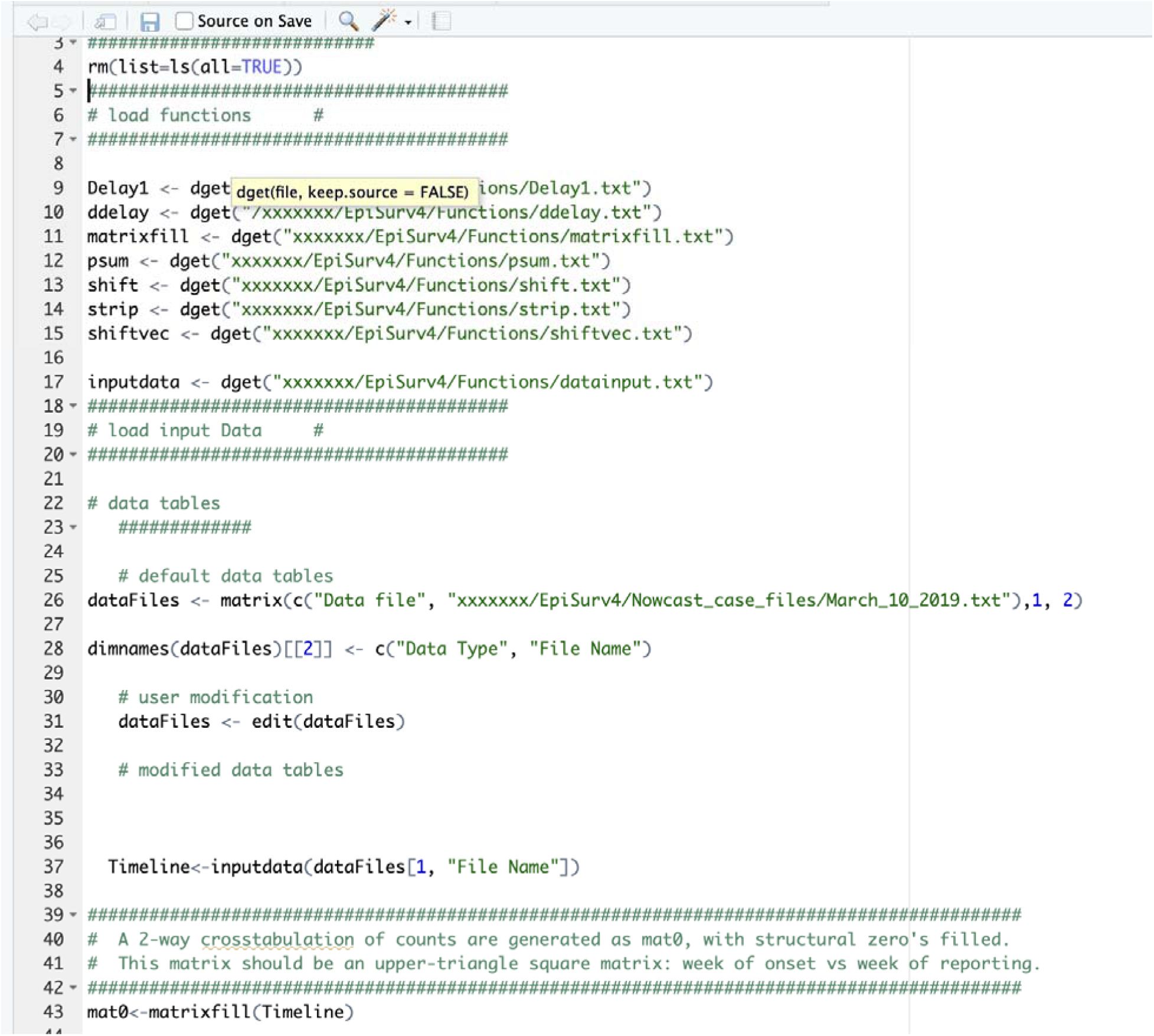
Components of the load-data-and-function.r file. Insert the pathway to the files and functions wher “xxxxxxx” is indicated.

In order to run the code to estimate the adjusted case incidence, open R console, type *Sourc (“c:/EpiSurv/load-data-and-functions.r”).* This will open a dialogue box. For the current data, type the input filename as: March_3_2019.txt. Then close the box (Fig 5).

**Fig 5.**
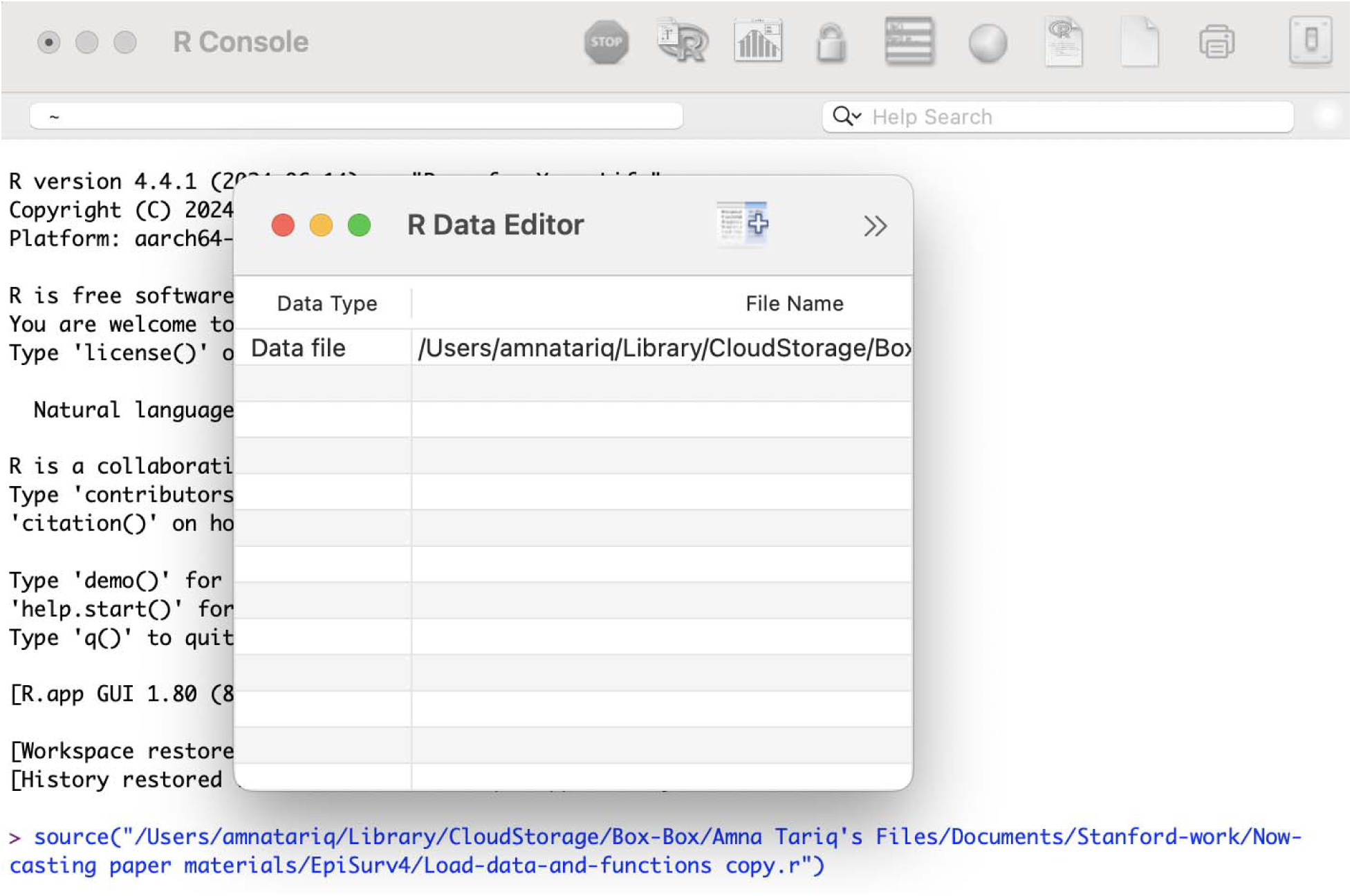
Dialogue box as seen using the code *Source (“c:/EpiSurv/load-data-and-functions.r”)* in R console.

In order to check the contents of the file, type ls(). It will produce a list of the functions and items that have been created. The following results are produced (Fig 6).

**Fig 6:**
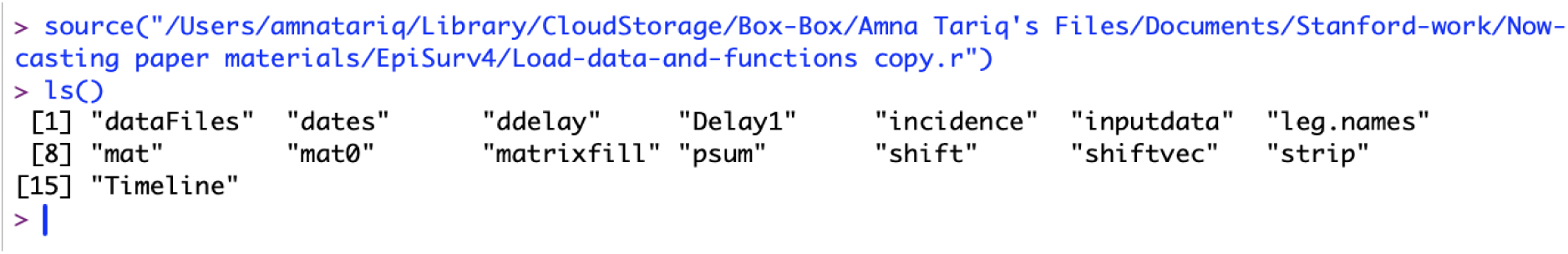
List of elements in ls().

## RESULTS

To check the components of the right and left upper triangular matrices, we provide the command “mat” and “mat0”. The matrix “mat” is an upper left triangular square matrix with rows representing the week of onset and columns representing the reporting delay (Fig 7). The matrix “mat0” is an upper right triangular square matrix with rows representing the week of onset and columns representing the week of reporting (Fig 8).

**Fig 7.**
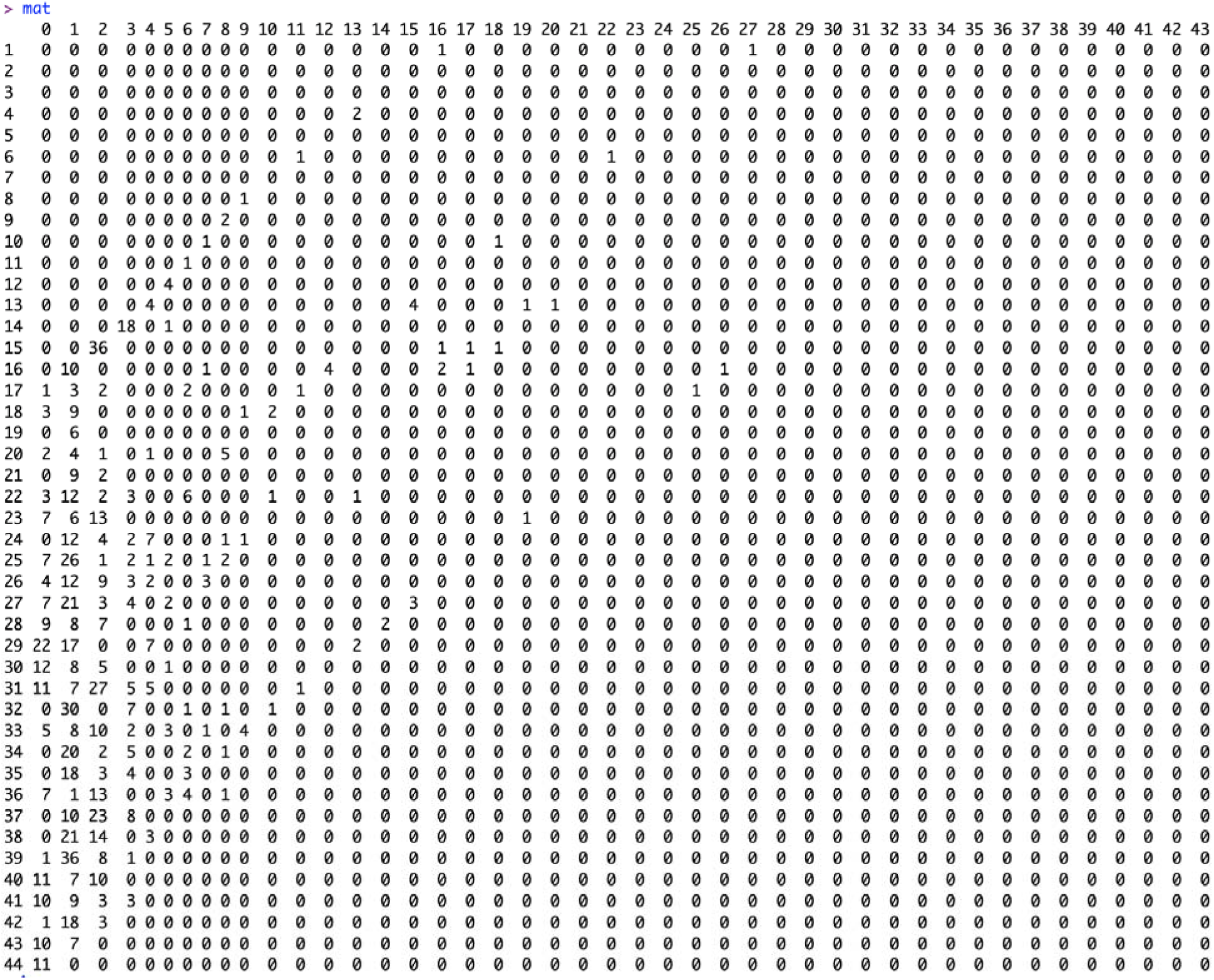
Elements of the matrix “mat” which is an upper left triangular square matrix with rows representing week of onset and column representing the reporting delay.

**Fig 8.**
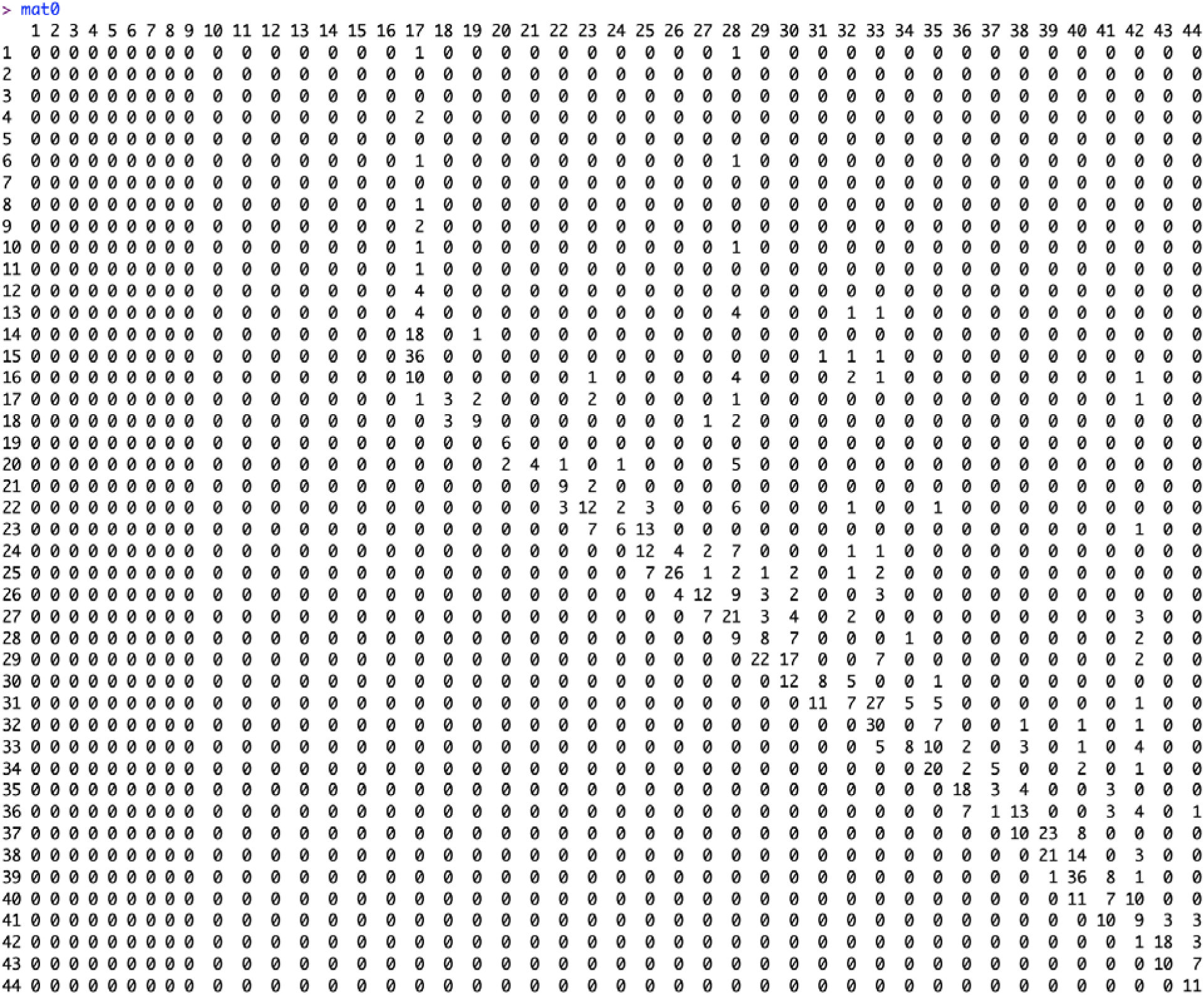
Elements of matrix “mat0” which is an upper right triangular square matrix with rows representing week of onset and column representing week of reporting.

The function “ddelay” contains the algorithms from Lawless (1994) [8]. The input data for the “ddelay” function is the matrix “mat”. For the “ddelay” function, there is a choice to provide the value of “m”. This allows us to select the most recent reporting period “m” (according to the week of reporting) that we believe is the time during which the reporting practice has been reasonably stable. If we set `m = ncol(mat)`, then the algorithm assumes a stable reporting delay distribution from the beginning of the dataset, which is not true in most instances. The command line to produce the required output is ***ddelay(mat, m).*** Fig 9 shows the results when m=7. The list of outputs it produces includes the estimated reverse-hazard with confidence limits, the right-truncated conditional probabilities as the delay-adjustment factor, and a table of reporting delay adjusted incidence (Figs 10, 11, 12).

**Fig 9.**
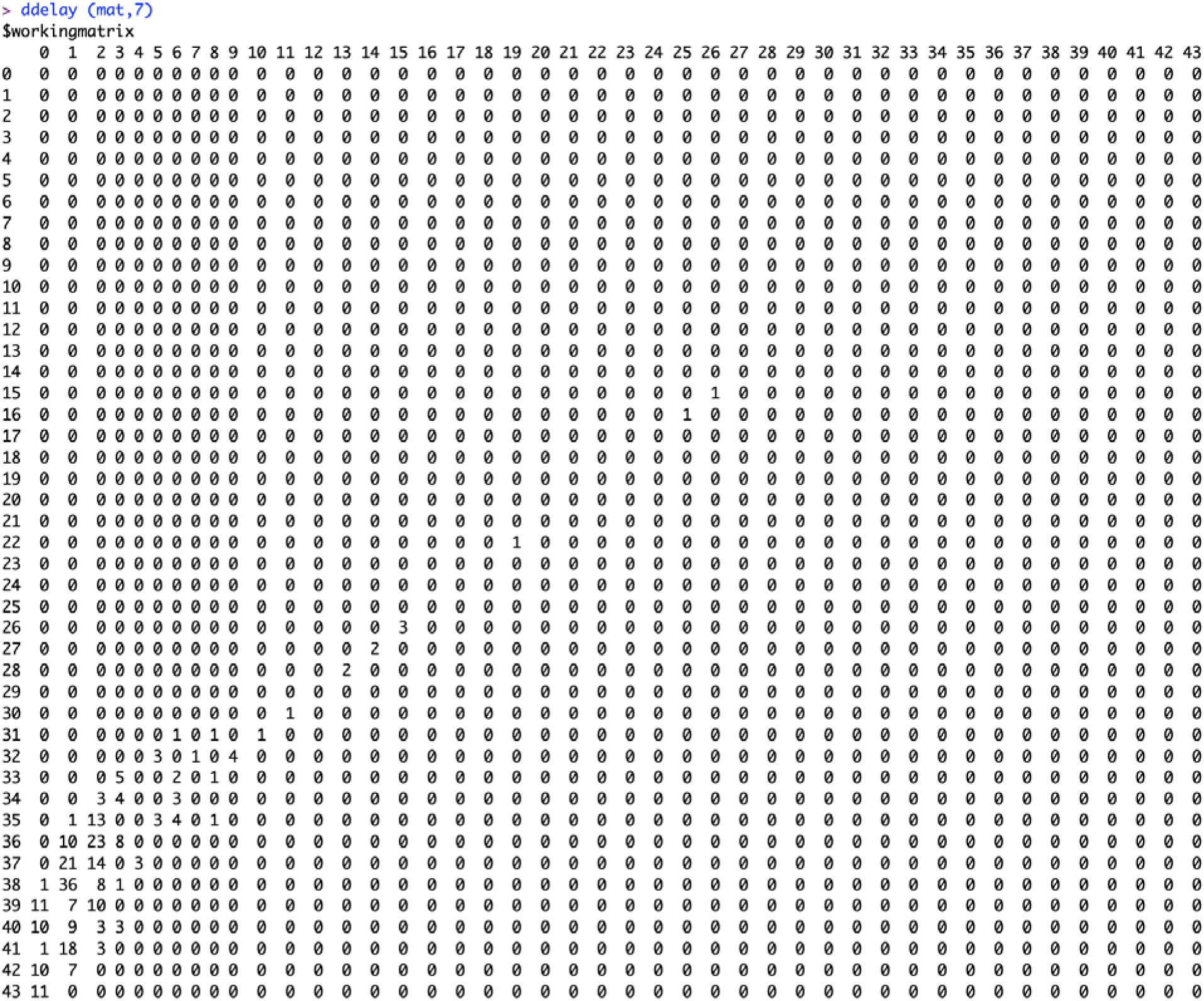
Elements of matrix “mat” with a non-stationary reporting delay window m=7.

**Fig 10.**
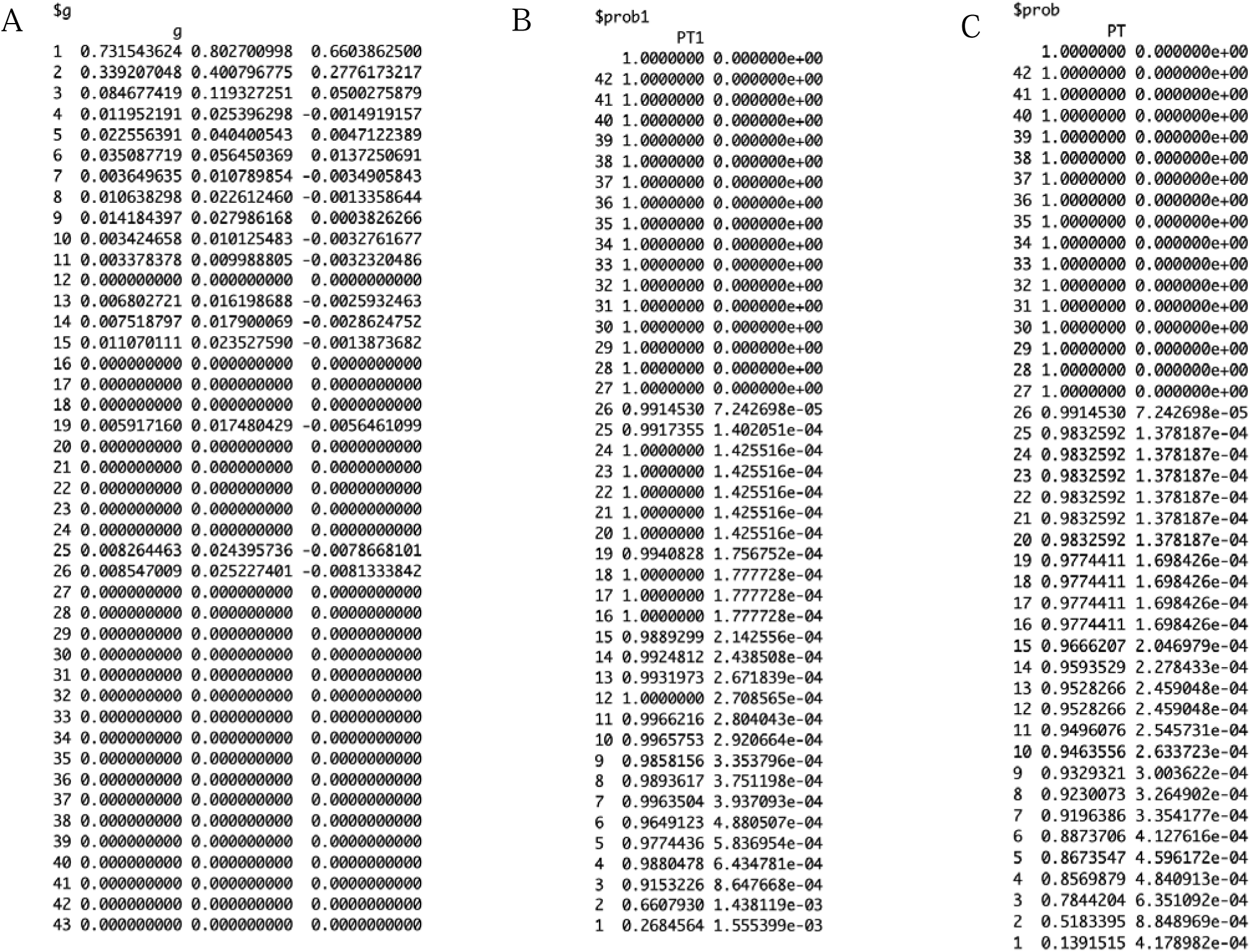
Probabilities for the data set generating the reporting delay adjusted incidence. The data set $g represents the estimated reverse hazards with the confidence intervals, $prob and $prob1 represent th right-truncated conditional probabilities as the delay-adjustment factor.

**Fig 11.**
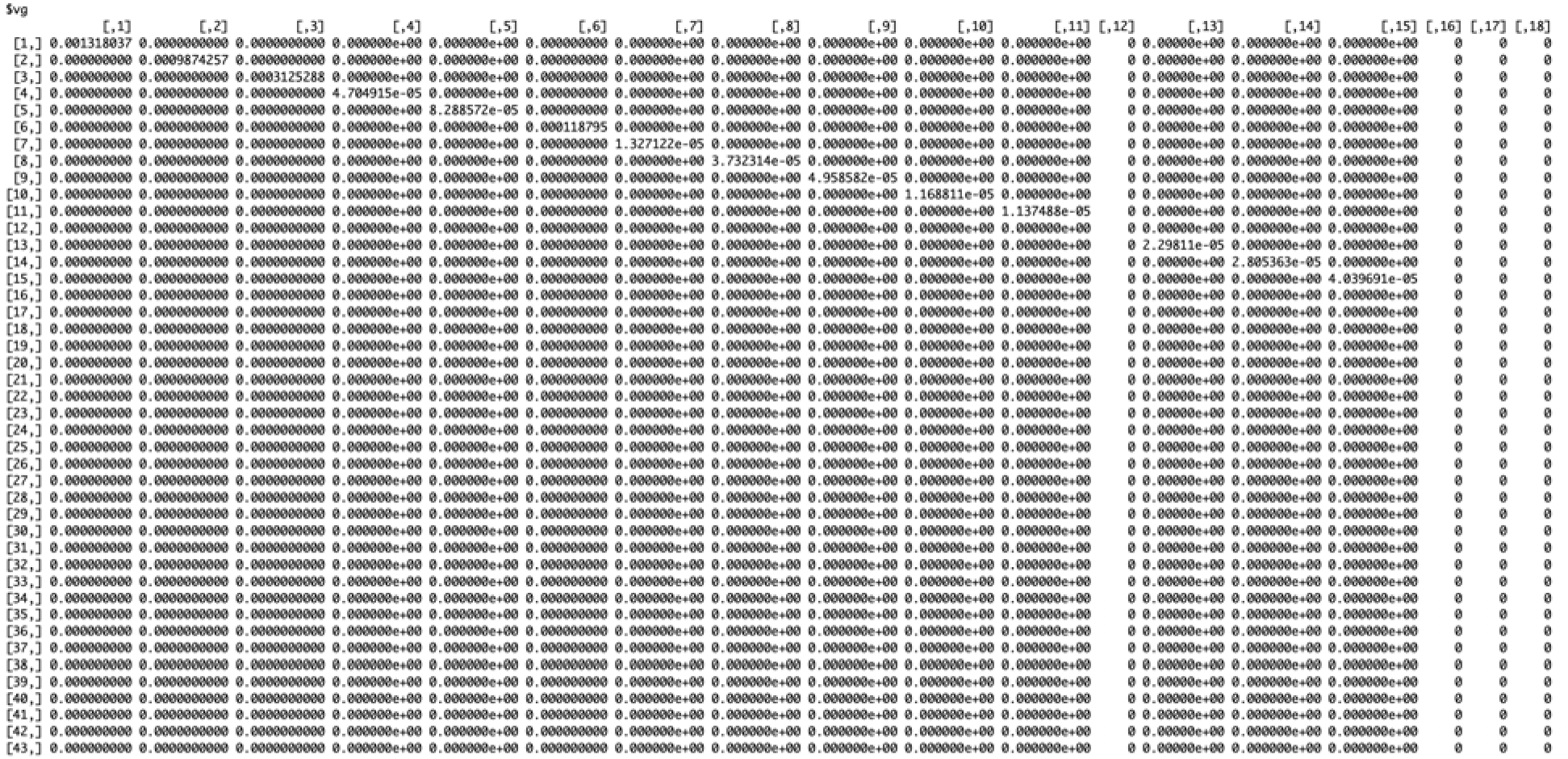
Matrix for probabilities for the week of onset versus the week of reporting.

**Fig 12.**
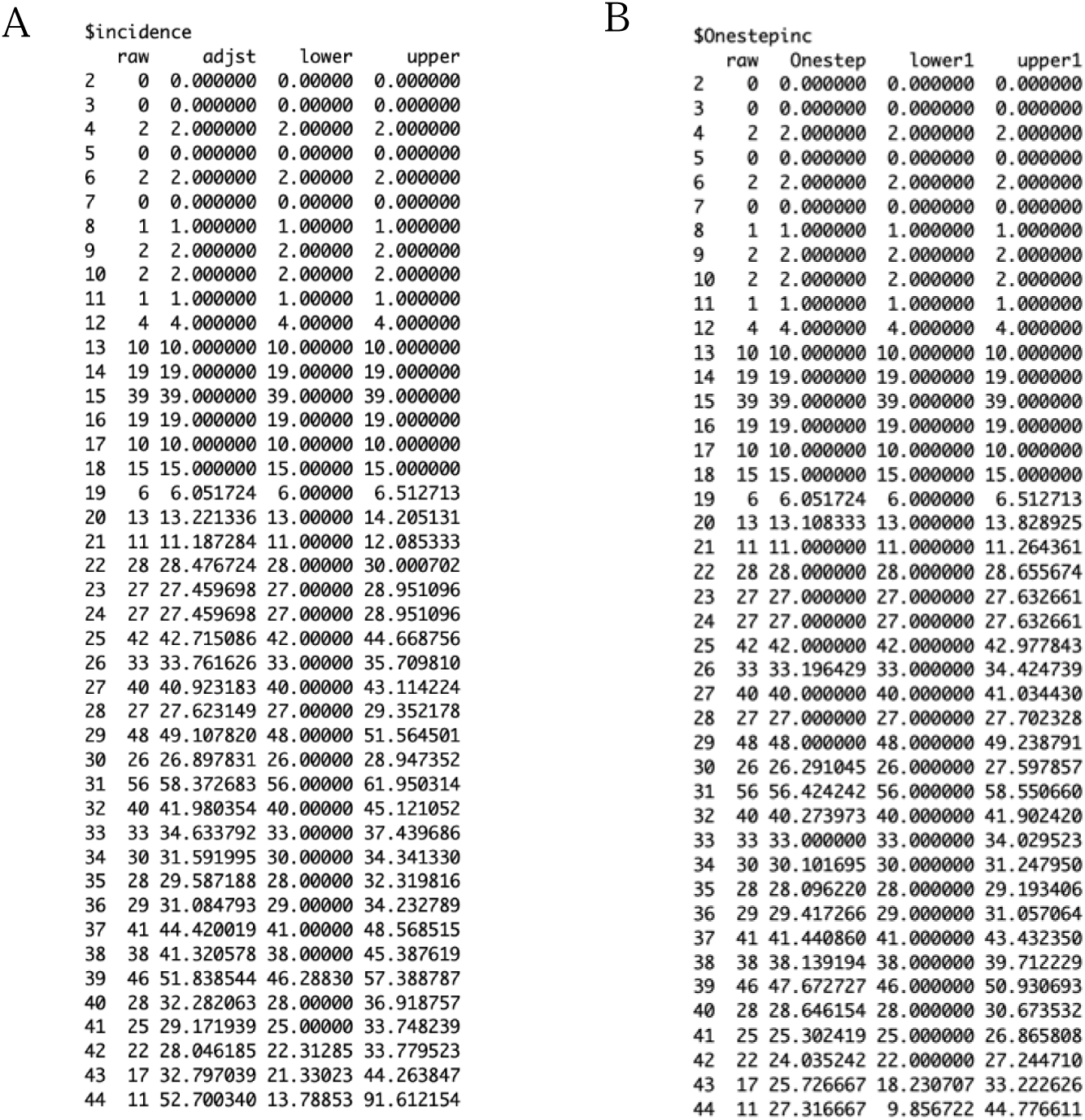
Panel A presented as $incidence contains the reporting delay adjusted incidence output with the first column representing the dates, second column representing the raw incidence data, third column representing the adjusted incidence, and fourth and fifth columns representing the upper and lower adjusted incidence, respectively. Panel B, presented as $Onestepinc, contains the one-step prediction of the data, with the first column representing the dates, the second column representing the raw incidence, the third column representing the one-step prediction, and the fourth and fifth columns representing the upper and lower adjusted one-step predicted incidence.

**Figure 13.**
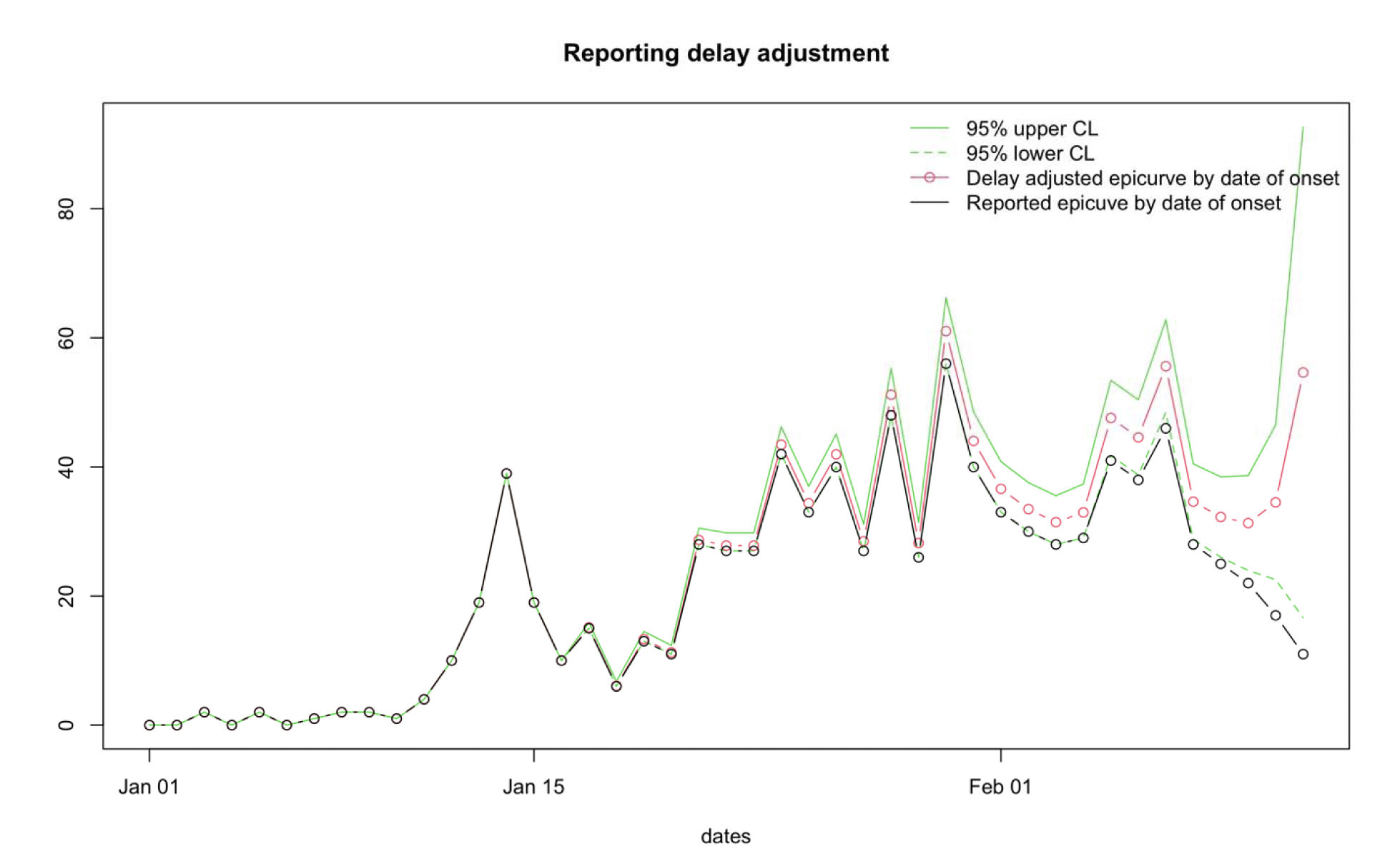
Reported delay adjusted incidence. Black line shows the raw incidence, green solid and dotted lines shows the upper and lower bounds of the adjusted incidence and red line with circles shows the adjusted incidence.

### Performance metrics

We assess the performance metrics of the different nowcasts by varying the value of “m” and comparing the difference from the true incidence, 8 weeks later (Figs 14, 15). We extract the reporting delay-adjusted incidence, true incidence, upper limit of the reporting delay-adjusted incidence, and the lower limit of the reporting delay-adjusted incidence from the output file (Fig. 12 Panel A) to estimate the performance metrics (Table 2).

**Fig 14.**
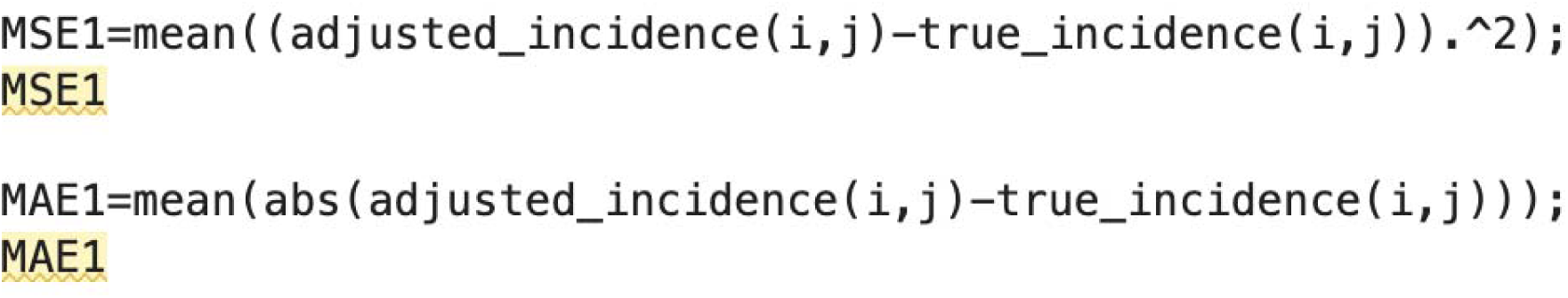
Matlab code to estimate the MAE (mean absolute error) and the MSE (mean squared error)

**Fig 15.**
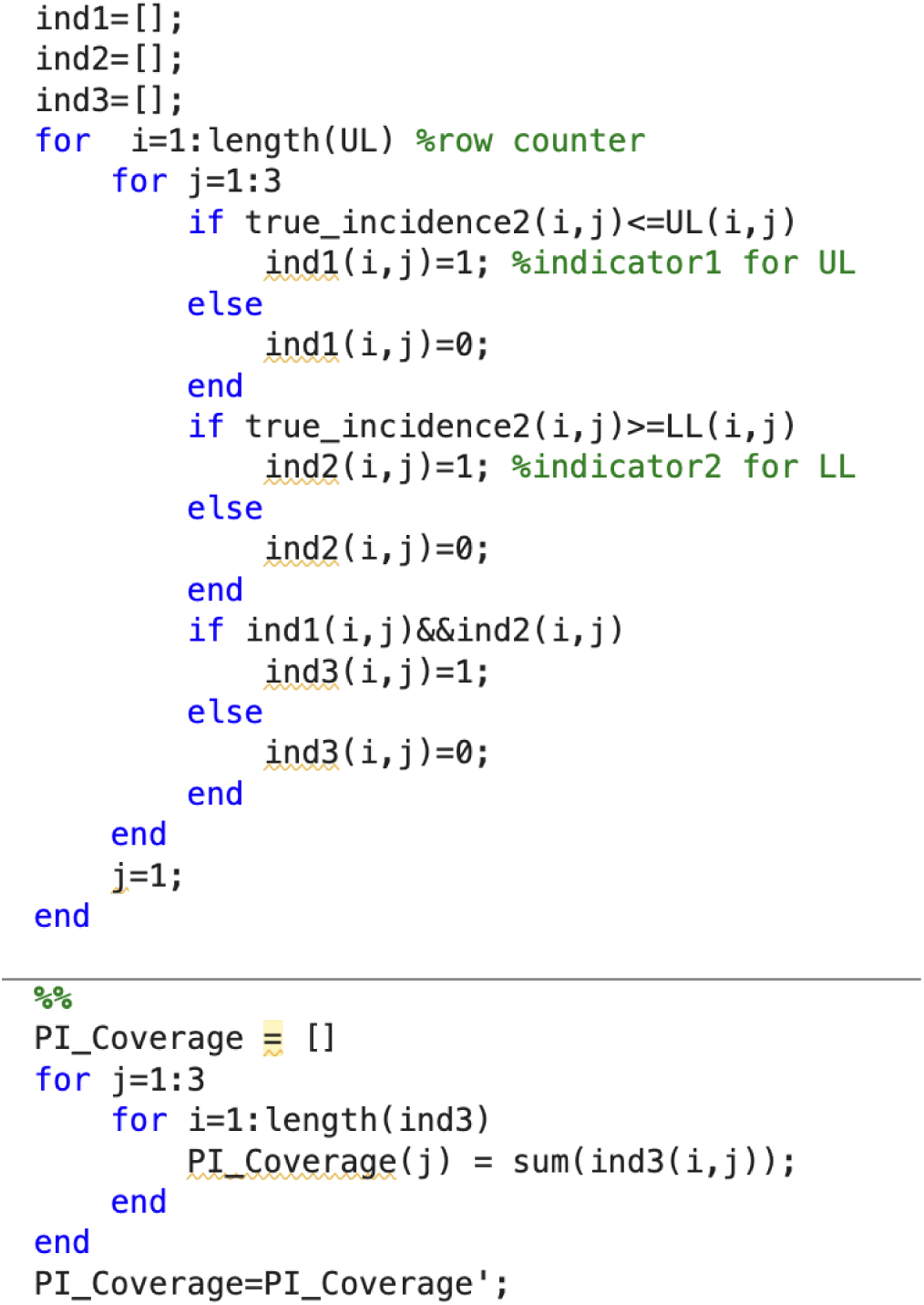
Matlab code to estimate the 95% PI coverage.

**Table 2.**
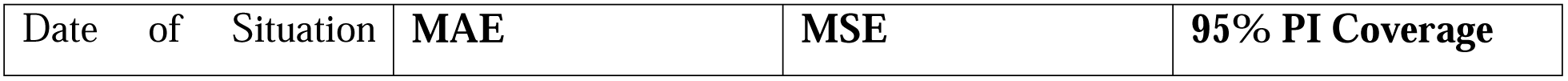
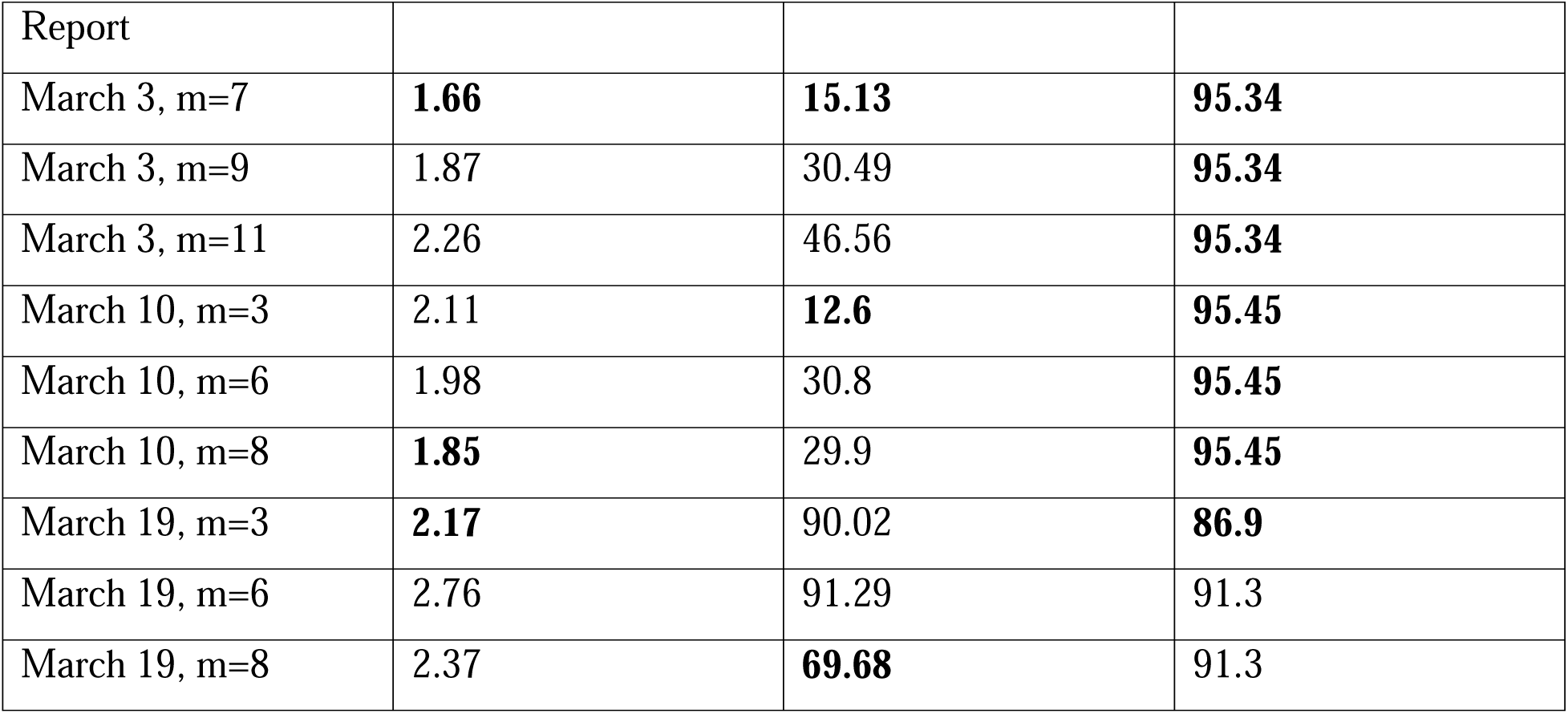
Results of performance metrics for selecting the best “m”. MSE represents the mean squared error, MAE represents the mean absolute error and 95% PI represents the 95% prediction interval coverage. Best value is presented in bold. The values of “m” can be varied according to the user. Any value of “m” can be applied and checked for its robustness using the performance metrics.

Based on the results of the performance metrics, the best “m” value would be m=7 for March 3, 2019, assuming a non-stationary reporting delay pattern of 7 weeks. Below, we show the panel figure representing the reporting delay adjusted curve using different values of “m” for the March 3, 2019, data file (Fig. 16).

**Fig 16.**
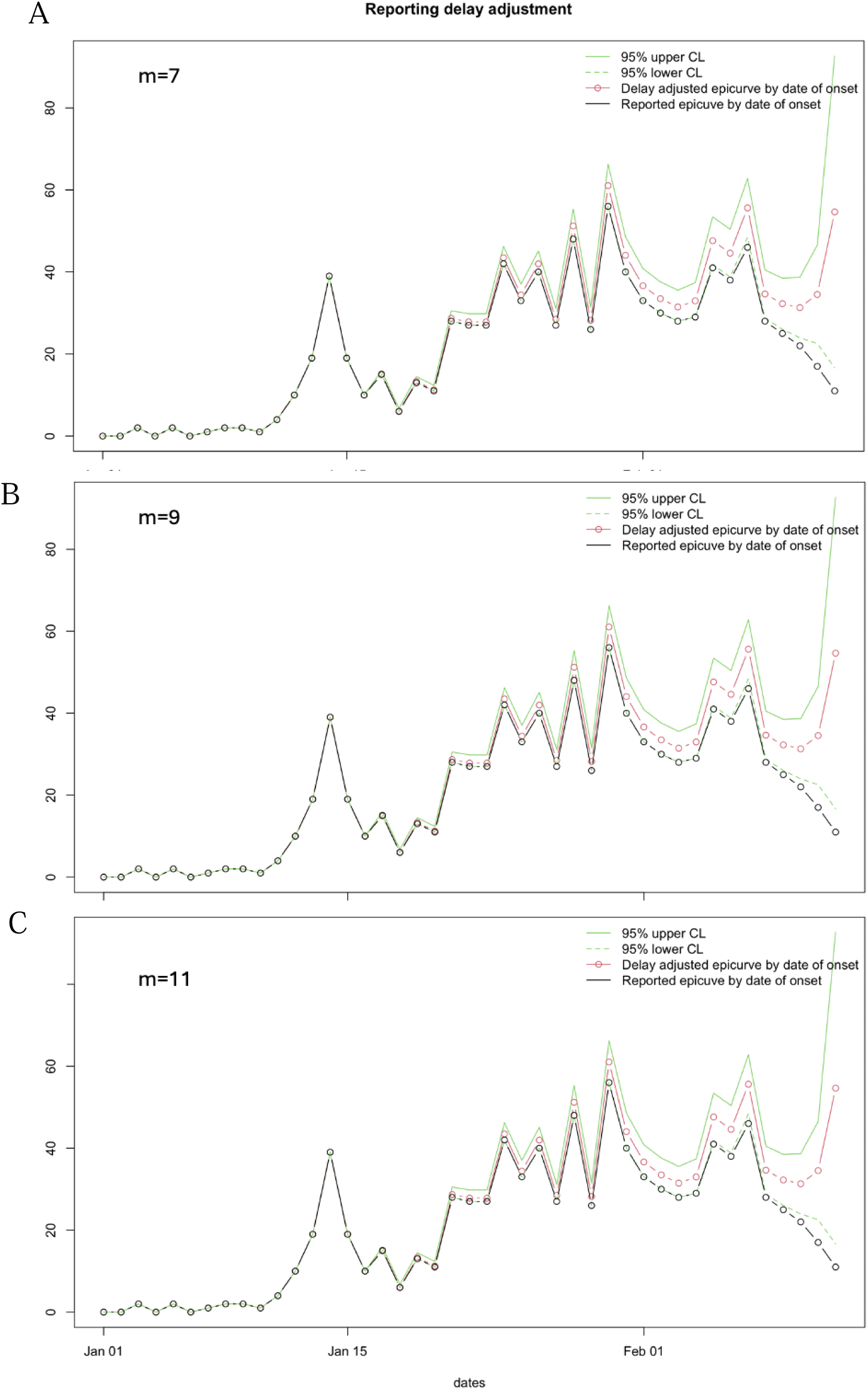
Reporting delay adjustment curves for the Ebola epidemic as reported on March 3, 2019, adjusted according to (Panel A), (Panel B) and (Panel C).

Similarly, when we use the data file for March 10, 2019, and “m” values of 3, 6 and 8, we get the following figure (Fig 17), and the performance metrics are given in Table 2. The window “m”=3 provides the best results for MSE (12.6) and the window of “m”=8 provides the lowest value of MAE (1.85). The 95% PI coverage is the same for all three values of “m”.

**Fig 17.**
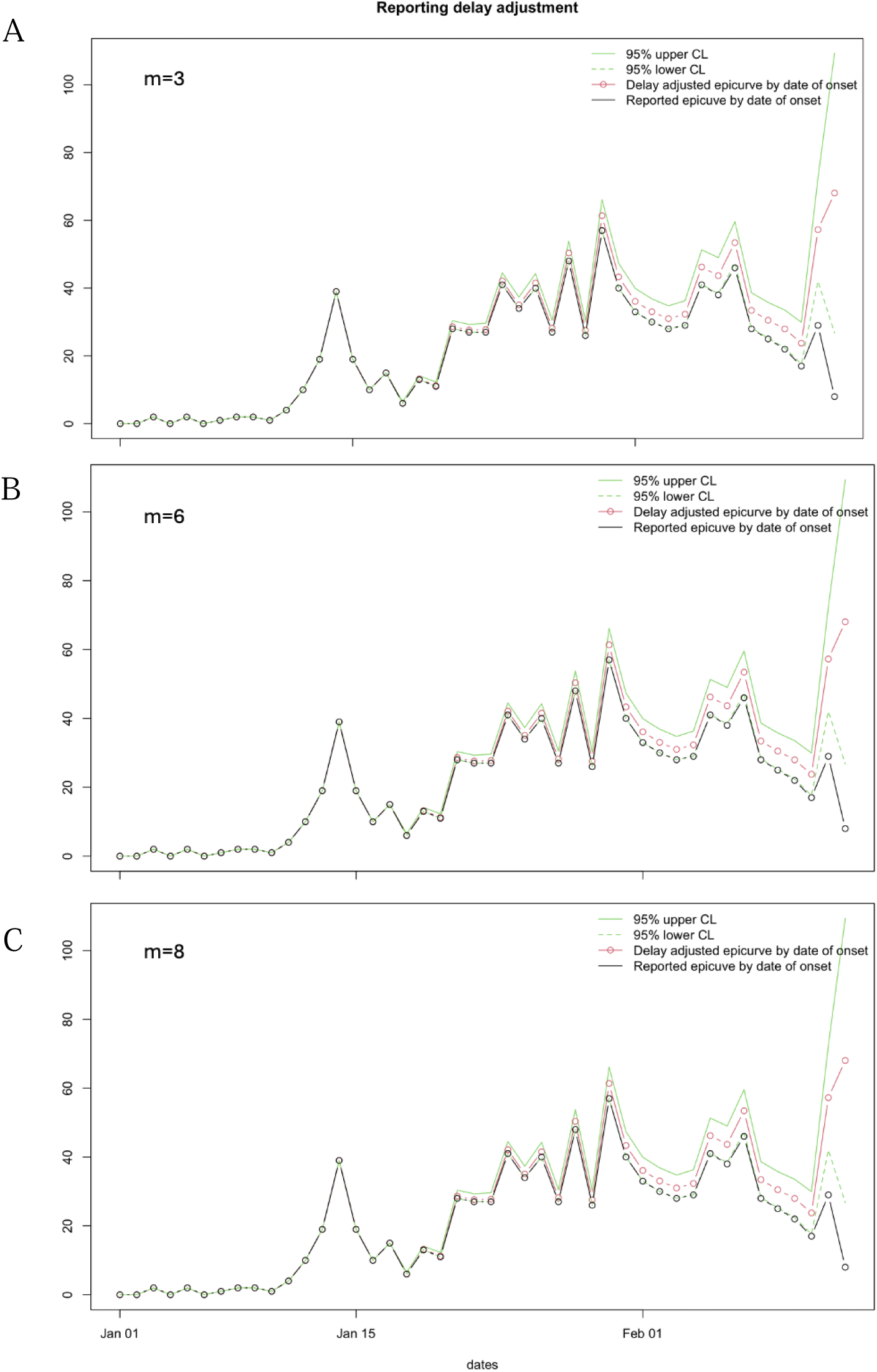
Reporting delay adjustment curves for the Ebola epidemic as reported on March 10, 2019, adjusted according to m=3 (Panel A), 6 (Panel B) and 8 (Panel C).

When we use the different values of “m” (3, 6, and 8) for March 19, 2019, the following figure can be produced (Fig. 18). The performance metrics are given in Table 2. Based on the results of the performance metrics, the window of “m”=3 provides the best results for the MAE (2.17) and “m”=8 provides the best results for MSE (69.68) and the 95% PI coverage (91.30).

**Fig 18.**
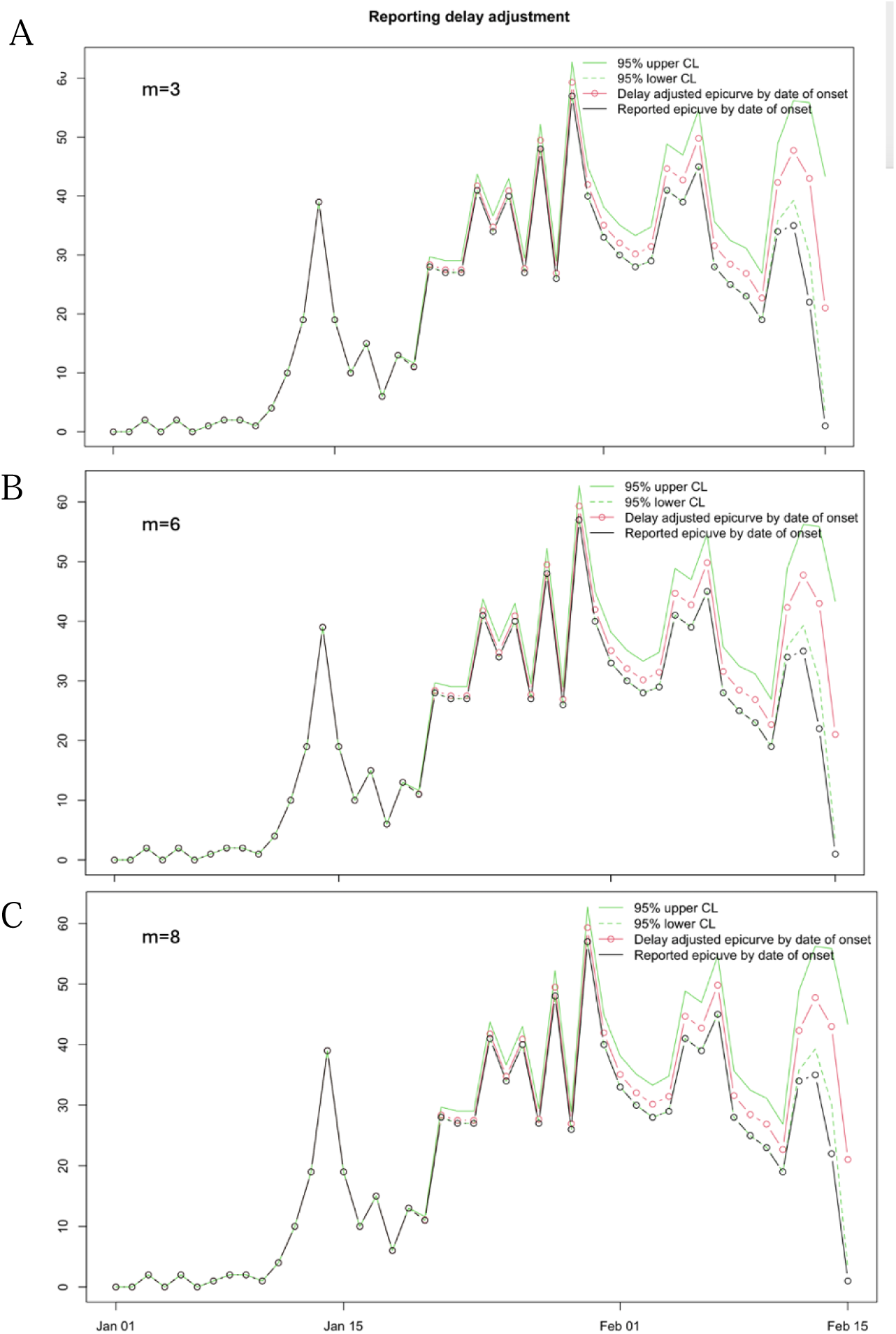
Reporting delay adjustment curves for the Ebola epidemic as reported on March 19, 2019, adjusted according to m=3 (Panel A), 6 (Panel B) and 8 (Panel C).

### Shiny app in R

Often, those training in public health or policymakers lack the programming expertise to quickly implement tools that can provide critical information for forecasting efforts, such as reporting-delay-adjusted curves. Therefore, we provide an R-Shiny App interface to facilitate the obtainment of reporting delay adjustment curves without requiring any previous programming or coding experience (Fig. 19).

**Fig 19.**
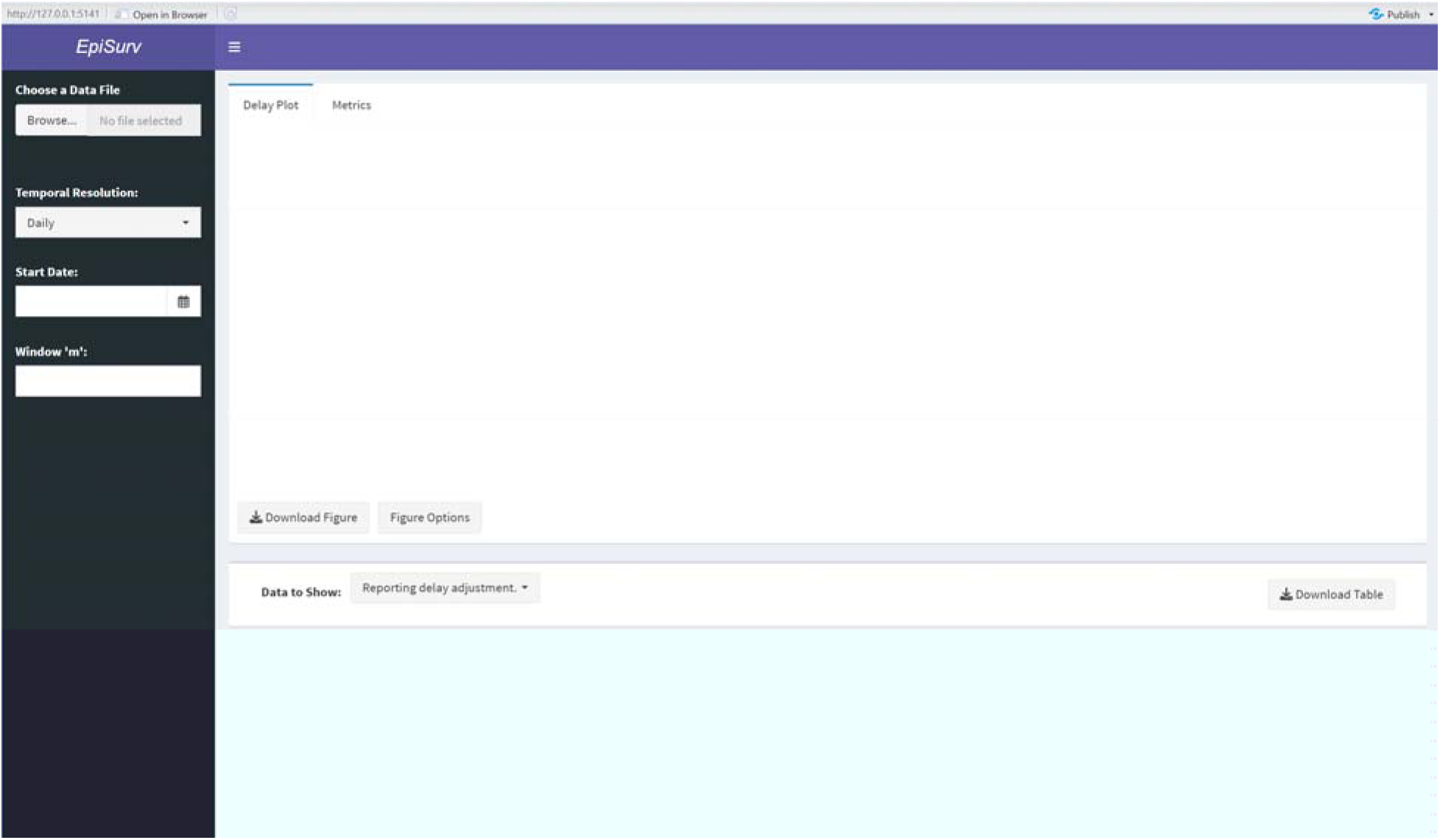
A screenshot of the *EpiSurv* R-Shiny App user interface. This figure provides a snapshot of the user interface for obtaining the reporting delay adjustment curves and other associated data files. As can be seen, the user has options to choose a data file, and then must specify information related to the temporal resolution of the data and the length of the reporting delay window, *m*.

The Shiny App interface utilizes the functionality discussed above but provides a graphical interface for users to input their data, specify the temporal resolution, and indicate the number of most recent time points (according to the reporting date) in which we believe the reporting practice has been reasonably stable (*m*). Additionally, users can customize and download the reporting delay adjustment curves, such as those shown in Fig. 13, obtain the data from Figs. 9-11 as ‘.csv’ files, and assess the reporting delay adjustment against the “truth” data. The following sections describe the implementation of the dashboard and provide additional examples of the output produced.

#### Installing and loading the Shiny App

As above, the Shiny App requires both R and RStudio for compilation. However, the implementation steps differ slightly:

- Download the ***EpiSurv*** folder containing the app and required functions from https://github.com/atariq2891/Reporting-delay-adjustment-code/tree/main.
- Load the **EpiSurv** project file in RStudio.
- Open the ‘app.R’ file, and click ‘Run App’.

The app requires multiple packages to function; however, they should be downloaded automatically upon the first rendering of the shiny application. Table 3 provides additional details regarding compilation requirements, as well as the web address for the host repository.

**Table 3.** *EpiSurv* metadata. Below includes details regarding the required software, versions, and packages needed to launch the EpiSurv R-Shiny application successfully. Additionally, it provides a link to the permanent repository, which contains all necessary functions, user interface files, and tutorial materials.

|  |  |
| --- | --- |
| **Require Software** | R ($\geq 4.3$), R-Studio (2024.09.0 Build 375) |
| **Compilation requirements<sup>1</sup>** | *pacman, shiny, shinydashboard, shinyWidgets, ggplot2,* |
|  | *DT, lubridate, shinyalert, chron* |
| **Permanent link to repository** | https://github.com/atariq2891/Reporting-delay-adjustment-code/tree/main/Shinyapp |

<sup>1</sup> Upon initial compiling of the dashboard, it checks if the required packages are downloaded. If they are not, R will proceed to install the packages for the given session. During this process, pop-up messages may appear asking if the user would like to compile the package. To successfully utilize all features of the dashboard, the user must select “yes”.

#### Inputting the data

The dashboard utilizes the same data format presented in Fig 2 and does not require any specific naming scheme. To load data into the dashboard, users can select the *Browse* button located under the “Choose a Data File” header and identify the file location within their personal computer. Once loaded, the user can then specify parameters related to the data’s temporal resolution and for calculating the adjusted incidence curve (Fig 20).

**Fig 20.**
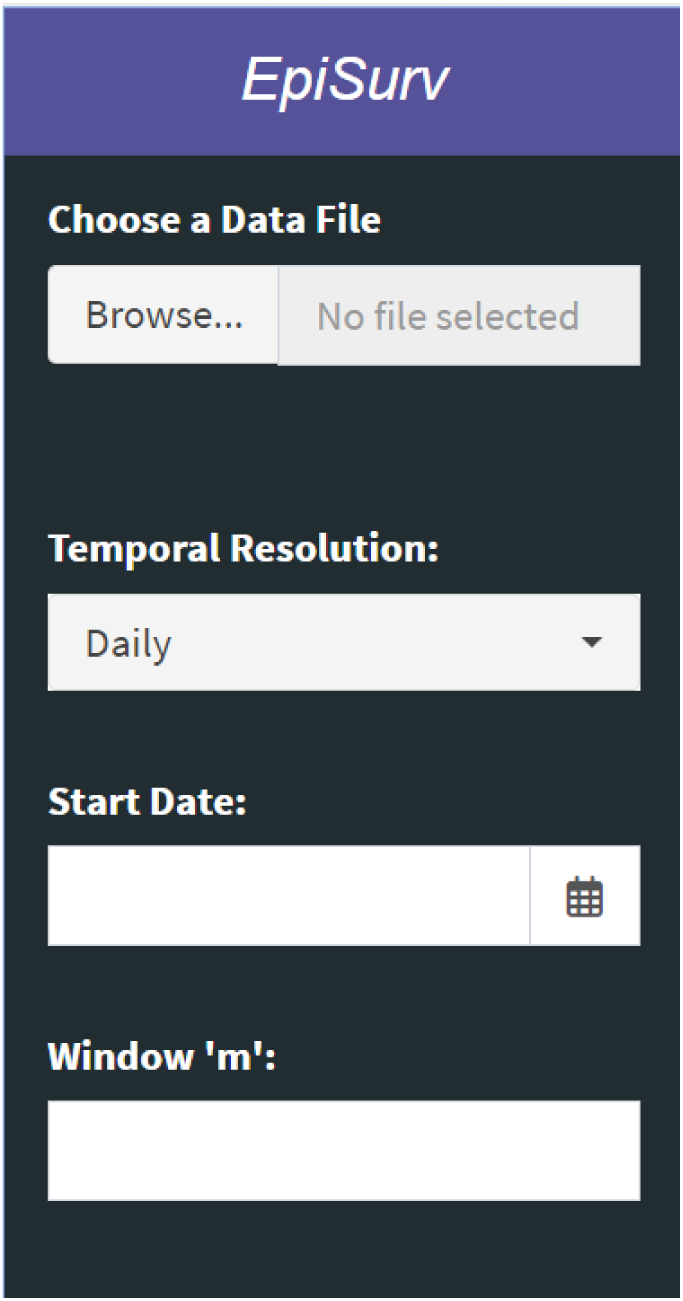
The user specifications available as part of the *EpiSurv* dashboard. Once the data ha been loaded, users must specify the temporal resolution of their data, the initial date included within the data, and the number of time points (according to reporting date) in which we believe the reporting practice has been reasonably stable (*m*).

#### User Specifications

Once data has been loaded, the users can then specify the temporal resolution of their data (i.e., *Temporal Resolution*). The dashboard works with continuous incident or mortality data for daily, weekly, monthly, and yearly periods. For each possible temporal resolution, a calendar will be available under *Start Date,* where users can select the first date (i.e., day, start of week, month, or year) of the available data. Only one date can be selected at a time. Users also have the option to adjust the value of *m*. When initially loading the data, the *m* parameter will default to the maximum number of time points available.

#### Available Output

Once data has been loaded, and a *Start Date* selected, the dashboard will auto-populate with the reporting delay adjustment curve (Figure 21), and multiple data sets including: (1) the reporting delay adjustment, (2) one-step ahead prediction of data, (3) estimated reverse-hazards with CI limits, (4) the right truncated probabilities, and (5) the matrix for probabilities.

**Fig 21.**
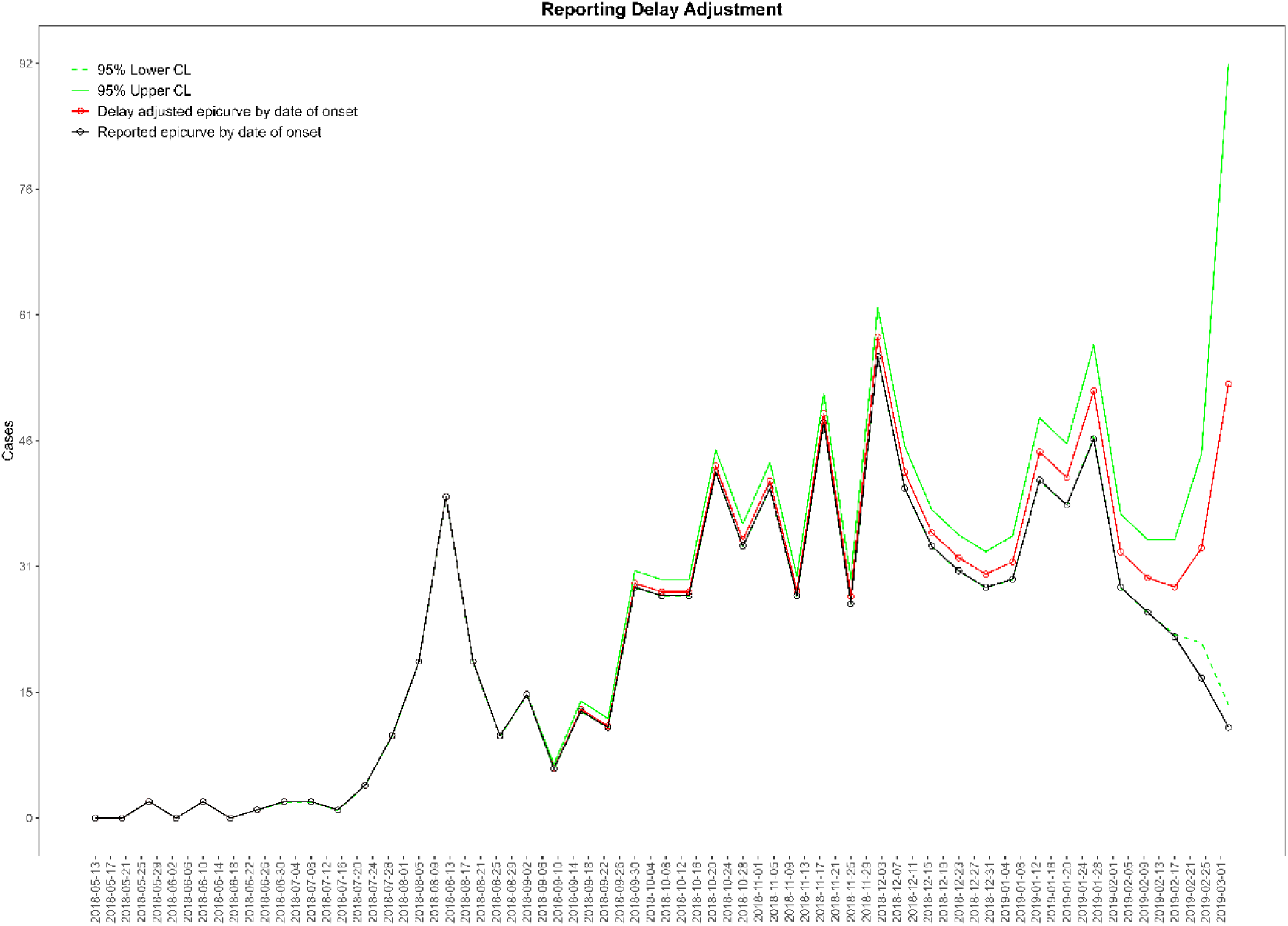
The reporting delay adjustment curve was obtained from the *EpiSurv* dashboard. This figure shows the reporting delay adjustment curve obtained from the *EpiSurv* dashboard using weekly Ebola data through May 3, 2019, and setting m = 7. The dashed green lines represent the lower 95% confidence limit, and the solid green lines are the upper 95% confidence limit. The red line represents the delay-adjusted epidemiologic curve by date of onset, and the black solid line and open circles represent the reported epidemiologic curve by date of onset.

The figure is fully customizable by clicking the *Figure Options* button and can be downloaded as a variety of file types. The data are available to download as *\*csv* files. When the underlying data, date options, or parameter *m* is changed, the files will update to reflect the new settings.

#### Performance Metrics

As discussed above, it is possible to assess the delay-adjusted incidence curve against data that is believed to represent the true incidence of disease over time. Once the reporting delay-adjusted curve has been calculated, the dashboard provides the mean absolute error (MAE), mean squared error (MSE), and 95% prediction interval coverage for download if the user provides the “truth” incidence data. The truth incidence data must be in either a *.txt* or *.csv* file format, containing only one column of counts over the time of interest and no column headers (Figure 22). The first row of the file must correspond to the first time point of data available in the delay-adjusted incidence curve. There is no required file naming scheme.

**Figure 22.**
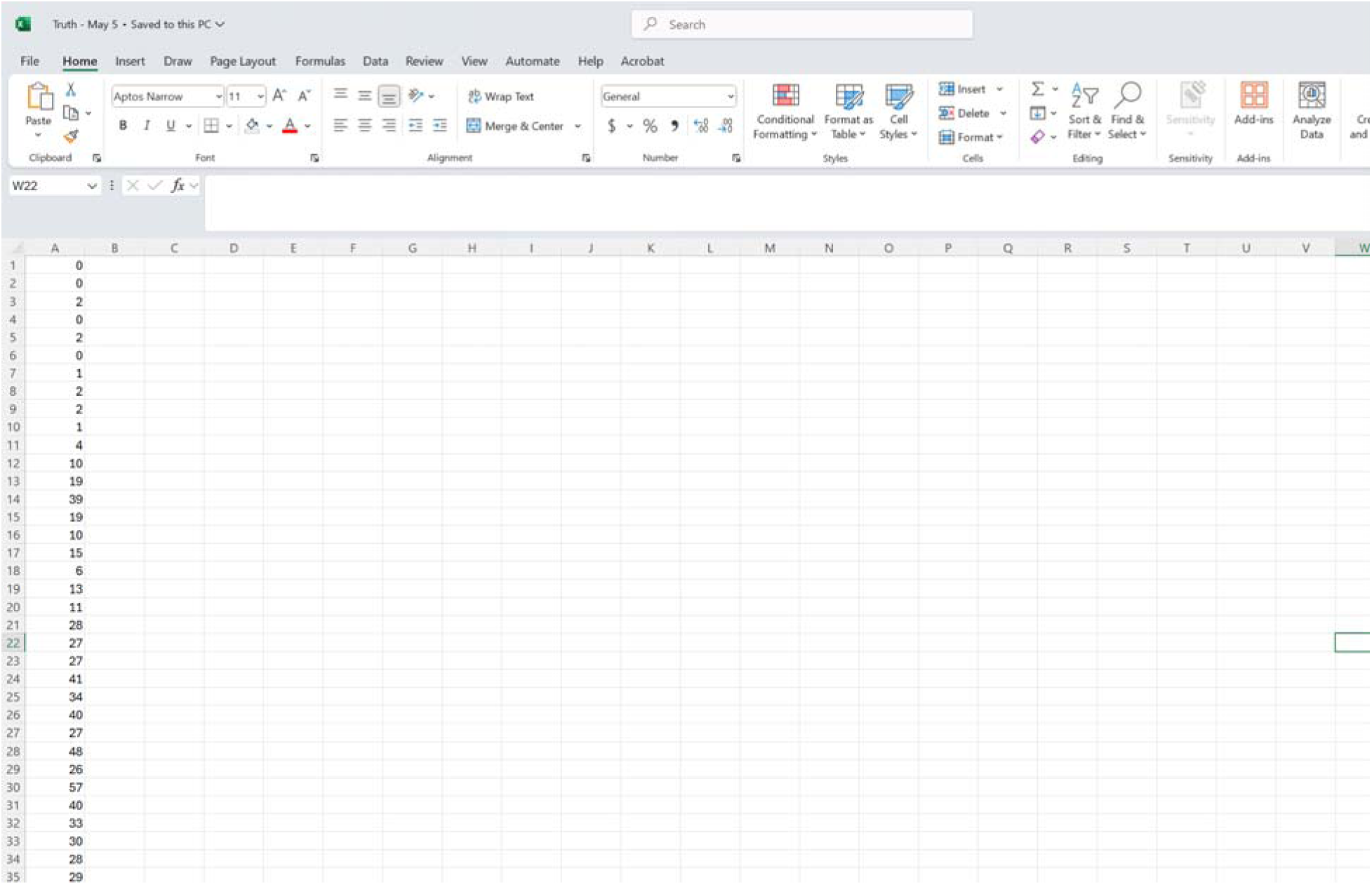
A screen shot of the required data format for the “truth” incidence data. The truth incidence data, used as part of the performance metrics calculations, must be a *\*txt* or *\*csv* file, and contain one column corresponding to the true incidence at each time point. The first row of data must correspond to the first time point available in the delay-adjusted incidence curve. There is no required naming scheme for the file.

The dashboard employs the same MAE, MSE, and 95% PI metrics provided above to ensure robust use with varying data types and lengths. Users can download the resulting metrics as a *\*csv* to their personal computer.

**Table 4.**
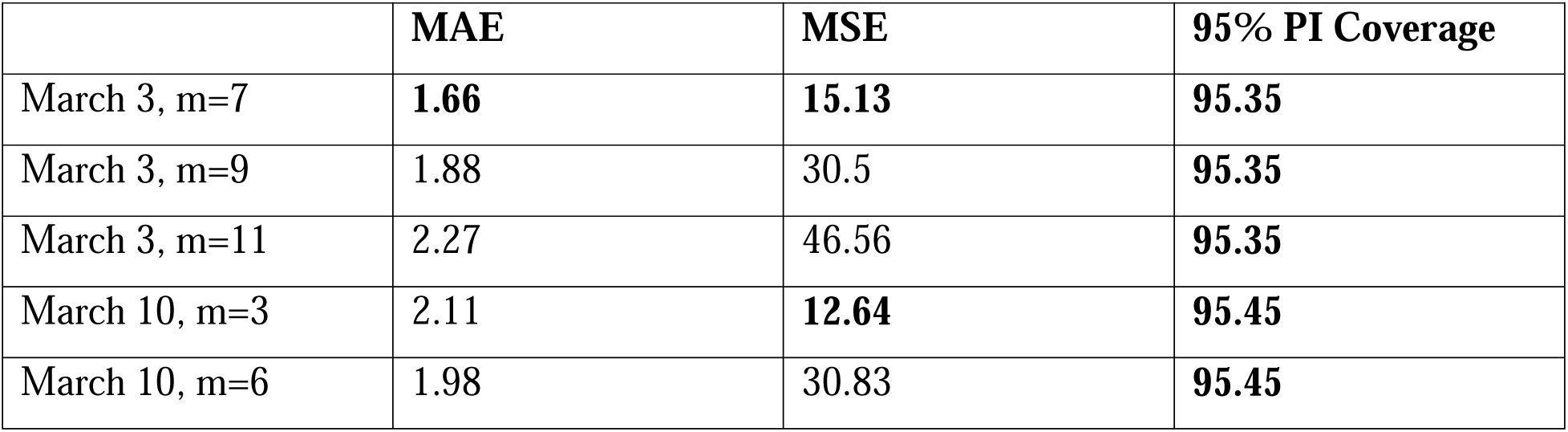
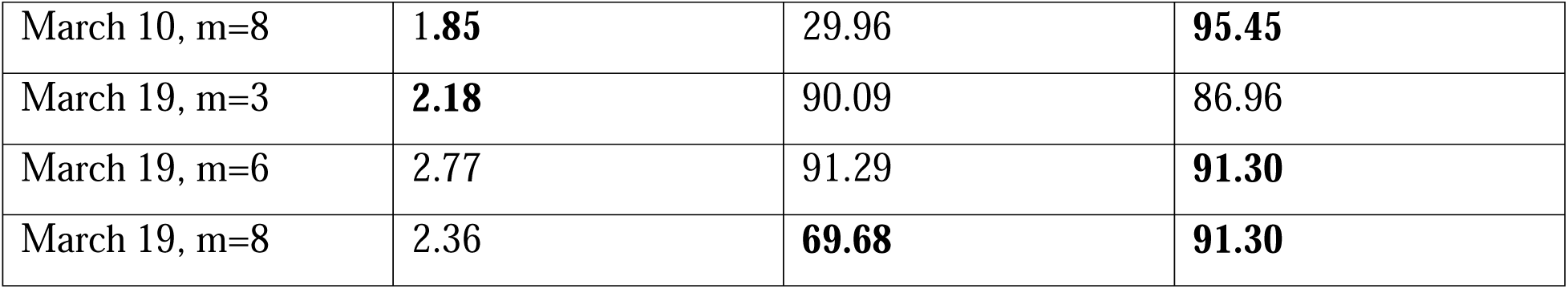
Results of performance metrics from the Shiny app. MSE represents the mean squared error, MAE represents the mean absolute error and 95% PI represents the 95% prediction interval coverage. Best value is presented in bold. The values of “m” can be varied according to the user. Any value of “m” can be applied and checked for its robustness using the performance metrics.

### Limitations to nowcasting the Ebola data

During real-time nowcasting of the Ebola epidemic in the DRC, we encountered two issues. First, there was an outstanding long delay that suddenly emerged as an outlier, as reported in the August 9, 2019, report. This long delay occurred possibly due to a case-review effort in one of the local reporting jurisdictions. Among the 104 cases reported during the most recent week (week 66), 33 cases were associated with onsets in the same week; 52 cases with onsets in the previous week; and 11 cases with onsets in Week 64. There were 4 cases with onsets in week 63, and 7 cases with long delays, including one case with onset in week 21 but reported in week 66 (reporting delay = 45 weeks). The result was a very rough pattern in the non-parametric estimate of the reverse hazard function along with overly estimated reporting delay adjustment. Normally, it would take a long time for the data to accumulate to determine whether this was just an outlier or a change in the reporting delay pattern (Fig 23).

**Fig 23:**
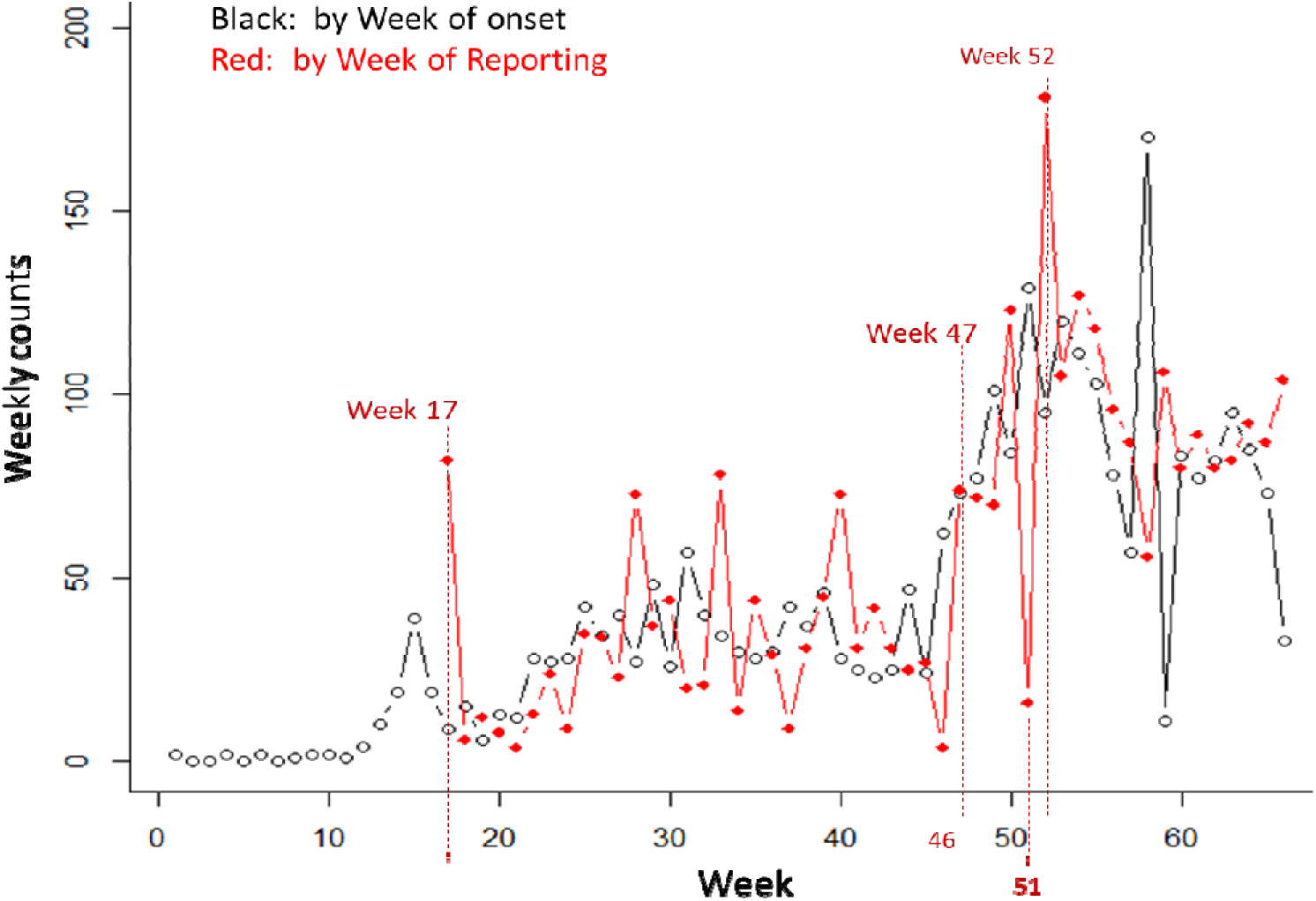
Depiction of the long reporting delays as reported in the situation report from August 9, 2019.

Secondly, in the situation report published on August 16, 2019, the number of Ebola cases increased from 33 cases (symptom onset week 66) to 67 cases (symptom onset week 67). This provided us with a very large value in the last reported case (that is n_{C0}), and the number of events in the current time point with delay = 0. This resulted in a sharp reporting delay-adjusted trend that cannot be easily deciphered by just looking at the figure. There is also no way of knowing if there was a sharp rise in the disease in the community or a sudden ramp-up of case reporting in the system. This kind of an error can be corrected by excluding the weeks before week 58 or by replacing the last data point in week 66 and week 67 as the running median of the previous 3 data points (Fig 24).

**Fig 24:**
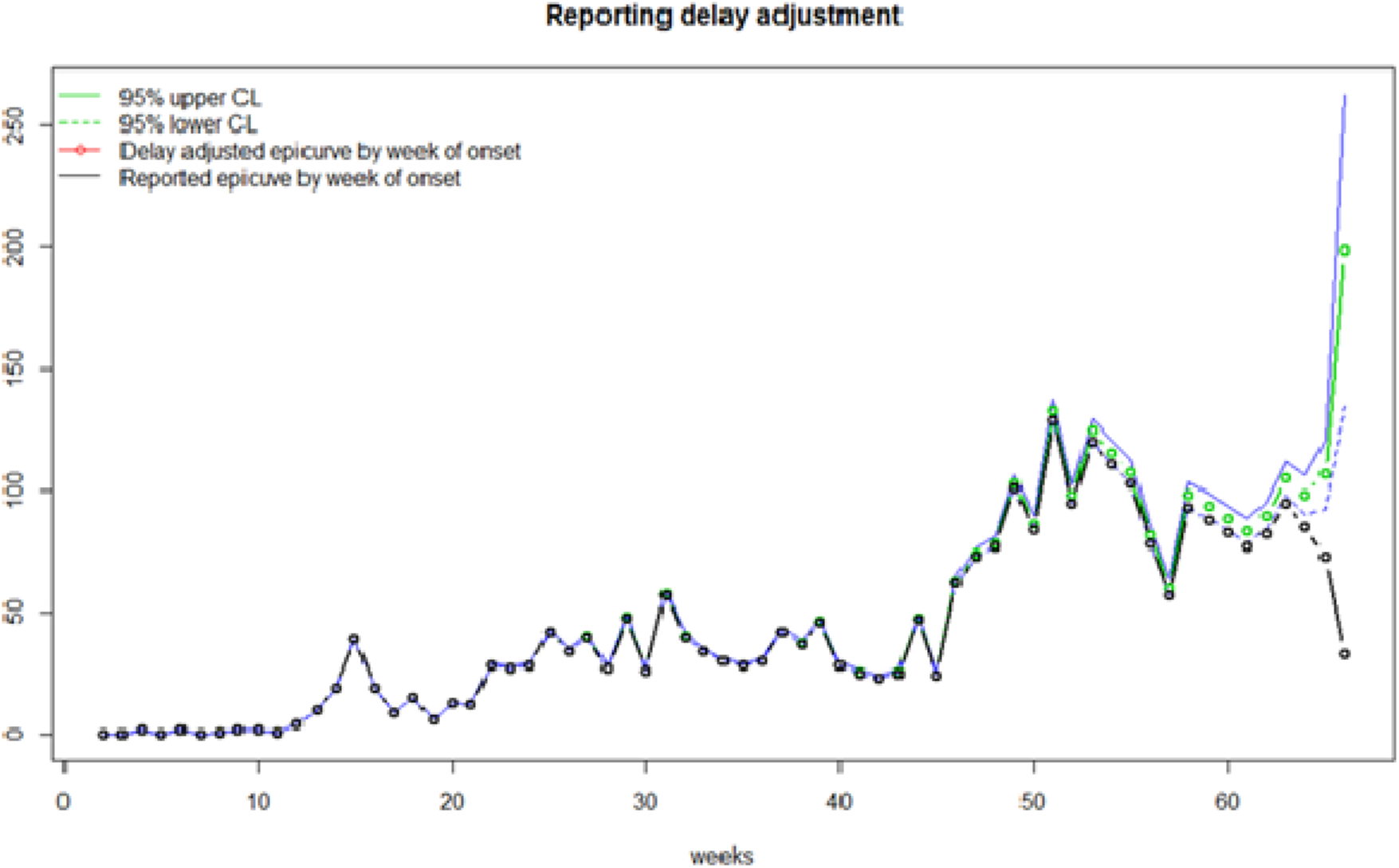
Sharp reporting delay adjustment in weeks 66 and 67 due to an increase in cases (33 to 67 cases) from week 66 to week 67 using the value of m=66 (entire dataset).

## Conclusion

This tutorial presents a practical nowcasting toolbox, Nowcast-It, designed to adjust for reporting delays in real time using a flexible frequentist approach. In particular, the method described in the toolbox has been frequently applied to nowcast the epidemic trajectories in near real-time [1, 4, 5]. The toolbox can be used as part of the curriculum of student training in mathematical biology, applied statistics, infectious disease modeling, and specialty courses in epidemic modeling and time-series nowcasting and forecasting. The data created via this approach can then be utilized to forecast the trajectory of the ongoing epidemics accurately. It is also a helpful resource for researchers and policymakers who can acquire better estimates of the missing and not reported cases that can be used to conduct short-term forecasts by relying on historical and real-time trajectory data of a process of interest.

The survival-based method implemented here is computationally efficient, does not require prior distributions or sampling algorithms, and offers intuitive parameter tuning through the selection of delay estimation windows. This makes it accessible for public health practitioners and researchers without deep expertise in Bayesian statistics or advanced computational methods. Moreover, the Nowcast-It toolbox includes built-in tools to evaluate model performance using metrics such as mean squared error, coverage probability, and prediction interval width— important features for evaluating forecast reliability.

Despite its strengths, the toolbox has several limitations. First, it relies on line list data, which may not always be available or complete, particularly in low-surveillance contexts. Second, while the toolbox provides point estimates and confidence intervals based on Lawless’ method, it does not offer full posterior distributions or simulation-based prediction intervals that are available in Bayesian approaches. In conclusion, Nowcast-It provides a practical and interpretable framework for correcting real-time reporting delays in infectious disease surveillance. Its combination of methodological rigor, computational efficiency, and accessibility makes it a valuable resource for epidemic response and training in applied infectious disease modeling.

## Author contributions

Conceptualization, G.C.; Methodology, G.C., P.Y.; Software, G.C., P.Y.; Validation, G.C., P.Y.; Formal Analysis, A.T.; Investigation, A.T, A.B.; Resources, G.C. .; Data Curation, A.T .; Writing – Original Draft Preparation, A.T., A.B.; Writing – Review & Editing, A.T. A.B. G.C.; Visualization, A.T., G.C.; Supervision, G.C.; Project Administration, G.C.; Funding Acquisition, G.C.

## Data Availability

The data and the code for this tutorial paper are publicly available on the Github repository https://github.com/atariq2891/Reporting-delay-adjustment-code/tree/main.

## Acknowledgements

NA

## References

1. Tariq A, Lee Y, Roosa K, Blumberg S, Yan P, Ma S, et al. Real-time monitoring the transmission potential of COVID-19 in Singapore, March 2020. BMC Medicine. 2020;18(1):166.10.1186/s12916-020-01615-9

2. McGough SF, Johansson MA, Lipsitch M, Menzies NA. Nowcasting by Bayesian Smoothing: A flexible, generalizable model for real-time epidemic tracking. PLoS Comput Biol. 2020;16(4):e1007735.10.1371/journal.pcbi.1007735

3. Van de Kassteele J, Eilers PHC, Wallinga J. Nowcasting the Number of New Symptomatic Cases During Infectious Disease Outbreaks Using Constrained P-spline Smoothing. Epidemiology. 2019;30(5)

4. Tariq A, Roosa K, Mizumoto K, Chowell G. Assessing reporting delays and the effective reproduction number: The Ebola epidemic in DRC, May 2018-January 2019. Epidemics. 2019;26:128–33.10.1016/j.epidem.2019.01.003

5. Munayco CV, Tariq A, Rothenberg R, Soto-Cabezas GG, Reyes MF, Valle A, et al. Early transmission dynamics of COVID-19 in a southern hemisphere setting: Lima-Peru: February 29(th)-March 30(th), 2020. medRxiv. 2020.10.1101/2020.04.30.20077594

6. Gebhardt MD, Neuenschwander BE, Zwahlen M. Adjusting AIDS incidence for non-stationary reporting delays: a necessity for country comparisons. Eur J Epidemiol. 1998;14(6):595–603.10.1023/a:1007406606892

7. G.C. Taylor. Claims Reserving In Non Life Insurance: Elsevier; 1985.

8. Lawless JF. Adjustments for Reporting Delays and the Prediction of Occurred but Not Reported Events. The Canadian Journal of Statistics / La Revue Canadienne de Statistique. 1994;22(1):15–31.10.2307/3315820

9. Brookmeyer R, Liao JG. The analysis of delays in disease reporting: methods and results for the acquired immunodeficiency syndrome. Am J Epidemiol. 1990;132(2):355–65.10.1093/oxfordjournals.aje.a115665

10. Kalbfleisch JD, Lawless JF. Estimating the incubation time distribution and expected number of cases of transfusion-associated acquired immune deficiency syndrome. Transfusion. 1989;29(8):672–6.10.1046/j.1537-2995.1989.29890020437.x

11. Roosa K, Tariq A, Yan P, Hyman JM, Chowell G. Multi-model forecasts of the ongoing Ebola epidemic in the Democratic Republic of Congo, March–October 2019. Journal of The Royal Society Interface. 2020;17(169):20200447.doi:10.1098/rsif.2020.0447

12. WHO. Ebola Situation Reports: Democratic Republic of Congo World Health Organization 2019 [Accessed on January 12, 2019].Available from: https://www.who.int/ebola/situation-reports/drc-2018/en/.

